# Profiling Muscle Mass, Function, and Molecular Signalling in Females Aged 18-80 and Their Associations with Sex Hormones

**DOI:** 10.1101/2025.07.21.25331955

**Authors:** Annabel J. Critchlow, Danielle Hiam, Steven J. O’Bryan, Ross M. Williams, Megan Soria, Viktor Engman, Karel van Belleghem, Ross P. Wohlgemuth, Andrew Garnham, Christopher S. Fry, David Scott, Séverine Lamon

## Abstract

Whether and how ovarian hormone fluctuations mediate the skeletal muscle response to ageing in females remains to be elucidated. We examined a tightly controlled, cross-sectional cohort of 96 females between 18-80 years of age to map the functional and molecular trajectory of muscle ageing and determine its relationship with female sex hormones. Across every decade, we quantified body composition (using dual-energy x-ray absorptiometry), muscle morphology (using peripheral quantitative computed tomography), and voluntary and evoked muscle function. Circulating sex hormone concentrations were measured with gas chromatography mass spectrometry and immunoassays. Morphology and gene expression of *vastus lateralis* muscle samples were assessed with immunohistochemical staining and RNA sequencing, respectively. Age was negatively associated with muscle mass, strength, and muscle fibre size, and positively associated with hybrid type I/II fibre prevalence and fibrosis. We found 37 unique patterns of gene expression across individual decades of age. Immune signalling, cellular adhesion, and extracellular matrix organisation pathways were the most upregulated with age, while mitochondrial function pathways were the most downregulated. Independently of age, circulating oestradiol and progesterone, but not testosterone, concentrations were positively associated with lean mass and negatively associated with hybrid muscle fibres across the lifespan. Oestrogen receptor binding sites were significantly enriched in upregulated genes in pre- versus post-menopausal muscle, suggesting a reduction in the translation of oestrogen target genes after menopause. Altogether, sex hormone fluctuations across the female lifespan may contribute to age-related muscle wasting, although longitudinal and interventional studies are needed to determine the causal nature of the relationship.

**Abstract figure:** 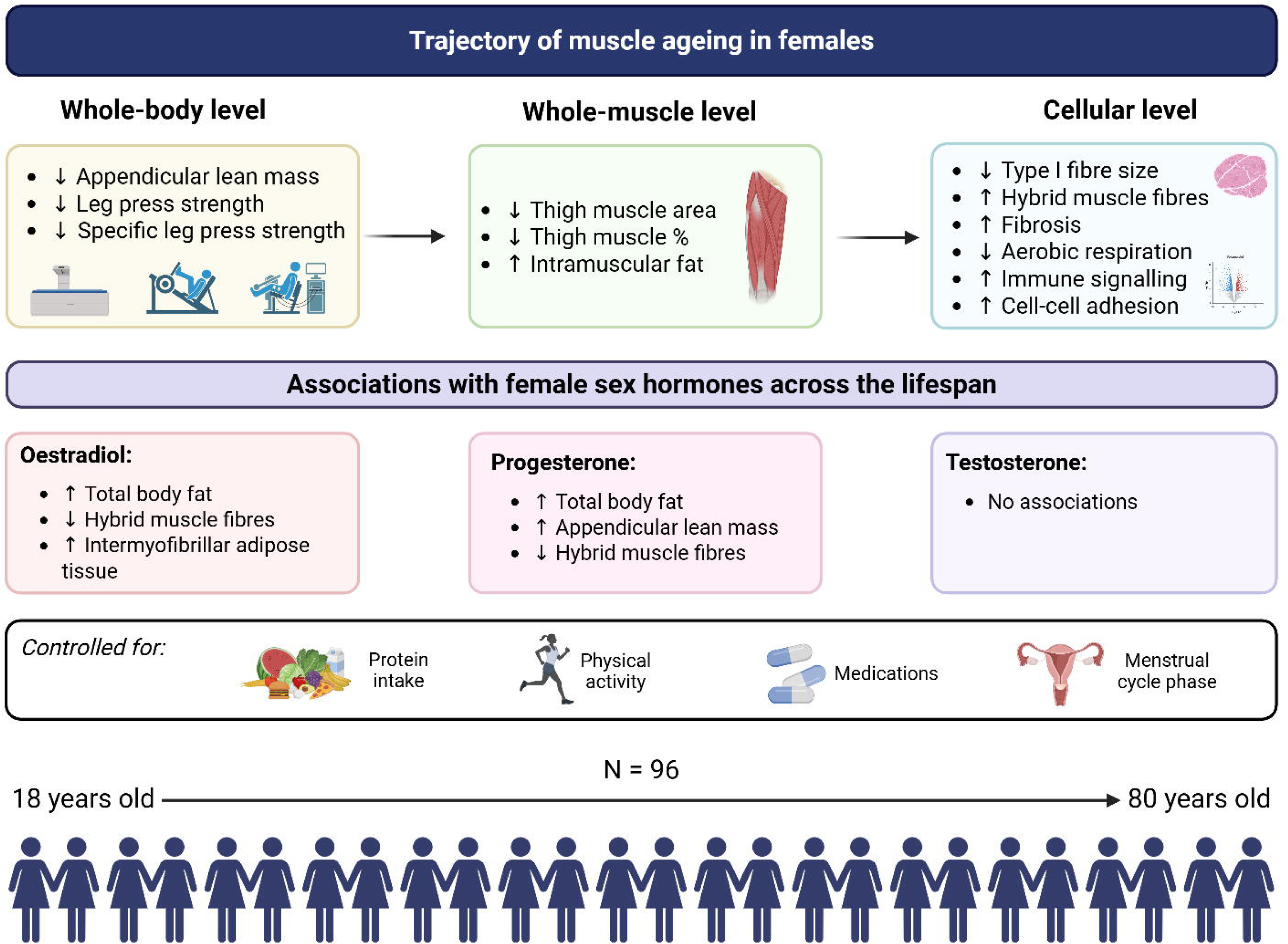

This study mapped the trajectory of muscle ageing at the whole-body, whole-muscle, and cellular level in 96 healthy females aged between 18 and 80 years old, while controlling for confounding lifestyle factors. Muscle mass and function declined with age, concomitant to a reduction in type I fibre size and increase in hybrid type I/IIa fibres. Patterns of muscle gene expression were mapped across ageing, showing an increase in immune cell signalling and a decline in mitochondrial respiration pathways. Circulating sex hormones were significantly associated with muscle mass, morphology, and gene expression across the lifespan.

**Key points summary**

1. Females live longer than males but experience worse disability in the later decades of life, highlighting the need to study female-specific patterns of ageing.
2. This study mapped female body composition, muscle morphology, function, and gene expression across every decade from 18 to 80 years of age in tightly controlled conditions and examined the relationships with circulating sex hormones.
3. Unique patterns of muscle gene expression across ageing showed an overall increase in immune signalling and a decrease in mitochondrial respiration pathways, but limited associations with circulating sex hormones.
4. Independently of age, circulating oestradiol and progesterone, but not testosterone, were associated with muscle mass and morphology across the lifespan, after adjusting for influential lifestyle factors (protein intake and physical activity).
5. Fluctuations in female sex hormones across the lifespan should be considered when developing therapies to mitigate age-related muscle wasting and improve the female health span.

## Introduction

Ageing is detrimental to skeletal muscle mass and function. Beyond the age of 40, muscle mass and strength decline by ∼8% and 15% per decade, respectively, with a rapid acceleration beyond the age of 70 (Kim & Choi, 2013). These impairments to skeletal muscle are underpinned by well-described molecular and cellular processes. Age-associated degradation of the neuromuscular junction causes denervation of muscle fibres and a loss of motor units, which can be followed by re-innervation by adjacent surviving motor units. Denervation leads to increased muscle proteolysis via the activation of atrophic signalling pathways, resulting in small, angular, and irregularly shaped fibres (Ehmsen & Höke, 2020). Other changes to muscle with age include increased oxidative stress, chronic inflammation and anabolic resistance, all of which lead to an overall deterioration of muscle cell quality and function (Bowen *et al*., 2015).

Despite skeletal muscle being a highly sex-specific tissue (Landen *et al*., 2023) with up to 3000 genes differentially expressed between human males and females at baseline (Oliva *et al*., 2020; Hanks *et al*., 2025), most of our understanding of skeletal muscle ageing originates from male human or animal models (James *et al*., 2023). A recent analysis of 478,438 adults over 40 years old from the UK BioBank however shows considerable differences in the muscle ageing trajectory between males and females (Fieldsend *et al*., 2025), where males exhibited a more pronounced loss of arm muscle mass, and females demonstrated a greater loss of muscle quality (i.e. force produced by a unit of muscle). At the molecular level, recent studies demonstrate prominent sex differences in the top ranked differentially expressed pathways in skeletal muscle from young to old individuals (De Jong *et al*., 2023), suggesting that biological sex may have an even greater influence than age on the resting muscle transcriptome (Pataky *et al*., 2023). These authors showed that 446 of the 503 (89%) genes that were differentially expressed between old (65-80 yr) male and female muscle were not located on sex chromosomes, suggesting they are regulated by other factors such as sex hormones.

Circulating sex hormones (e.g. oestrogens and androgens) are a major factor underlying systemic sex differences in mammals (Blencowe *et al*., 2022). The main androgenic and oestrogenic sex hormones, testosterone and oestradiol (E2, the major form of oestrogen), respectively, can act on skeletal muscle by binding to their specific receptor (Wiik *et al*., 2009), triggering their translocation to the nucleus and transcription of their specific target genes (Heldring *et al*., 2007). Both males and females produce E2 and testosterone, but their profiles vary between sexes and across the lifespan. Across the female lifespan, circulating E2 levels can peak at up to ∼1700pmol/L during ovulation and fall to ∼70pmol/L after the 5^th^ decade of life, marking the onset of menopause and the cessation of E2 production from the ovaries (Frederiksen *et al*., 2020). There are evident associations between declining oestrogen levels and deterioration of muscle mass and function (Critchlow *et al*., 2023, 2025) but distinguishing between the effects of age and oestrogen deficiency is difficult in humans as these processes occur simultaneously. We recently reported that, independent of age, total and free circulating E2 is positively associated with muscle mass, and the decline in free E2 over ∼5 years is associated with a loss of handgrip strength (Critchlow *et al*., 2025).

Despite these population-based results, the effects of the transition from high to low circulating E2 on the composition and functioning of skeletal muscle are not fully understood. Many animal studies have adopted ovariectomised rodent models to mimic and investigate the causal effect of oestrogen deficiency on skeletal muscle, confirming that the cessation of ovarian hormone production causes significant detriments to muscle composition, function, and metabolism, which may be rescued with subsequent E2 treatment (Pellegrino *et al*., 2022). Specific target pathways of E2 action in skeletal muscle may include mitochondrial function, satellite cell number and activation, protein synthesis and degradation, and the muscle damage and regeneration response (McClung *et al*., 2006; Barbosa *et al*., 2016; Larson *et al*., 2020; Cho *et al*., 2021). While useful to establish cause and effects relationships, rodent models come with several limitations warranting further investigations in tightly controlled human cohorts. Firstly, the invasive removal of the ovaries causes sudden cessation of ovarian hormone production, as opposed to the gradual decline experienced across several years of the menopausal transition. Secondly, the frequent use of young animals (Barbosa *et al*., 2016; Cho *et al*., 2021) ignores the context of an aged muscle phenotype, therefore omitting the influence of major factors that heavily impact the cellular environment, such as oxidative stress and inflammation (Mansouri *et al*., 2006; Franceschi & Campisi, 2014). Together these factors limit the direct translation of findings from rodents to human populations.

Few human studies have comprehensively investigated the effects of ageing, especially as a continuous variable, on female skeletal muscle, and even fewer have investigated the effect of the menopausal transition, the findings of which are currently inconclusive (Critchlow *et al*., 2023). Therefore, the aims of this study are two-fold: 1) to map body composition, muscle morphology, function, and gene expression in a tightly controlled cohort of 96 females from 18-80 years of age, and 2) to determine the associations of these outcomes with the dynamic changes of circulating sex hormone concentrations occurring across the lifespan.

## Methods

### Participants

#### Study outline

Ethical approval was granted by Deakin University Human Research Ethics Committee (project number 2021-307). Ninety-six females between the age of 18 and 80 were recruited for this study, with each decade of age represented in the cohort. All participants were provided with a plain language statement and gave written informed consent prior to participation, which conformed to the standards set by the Declaration of Helsinki (2013) and its later amendments, except for registration in a database. Inclusion criteria included healthy biological females with a body mass index (BMI) between 18-35kg/m^2^ who were free from cancer, neurological disease or any other condition that would prevent them from performing the tests safely and were not currently pregnant or lactating. Menopausal status and use of hormonal contraception or menopausal hormone therapy were defined during the medical screening phase. Postmenopausal status was informed by self-reported cessation of menstrual periods for >12 months and the presence of typical postmenopausal symptoms (Jane & Davis, 2014). Participants were recruited from the local community via Facebook advertisements, the Deakin Institute for Physical Activity and Nutrition website, posters, and word of mouth. An outline of the study timeline is shown in Figure 1.

**Figure 1.**
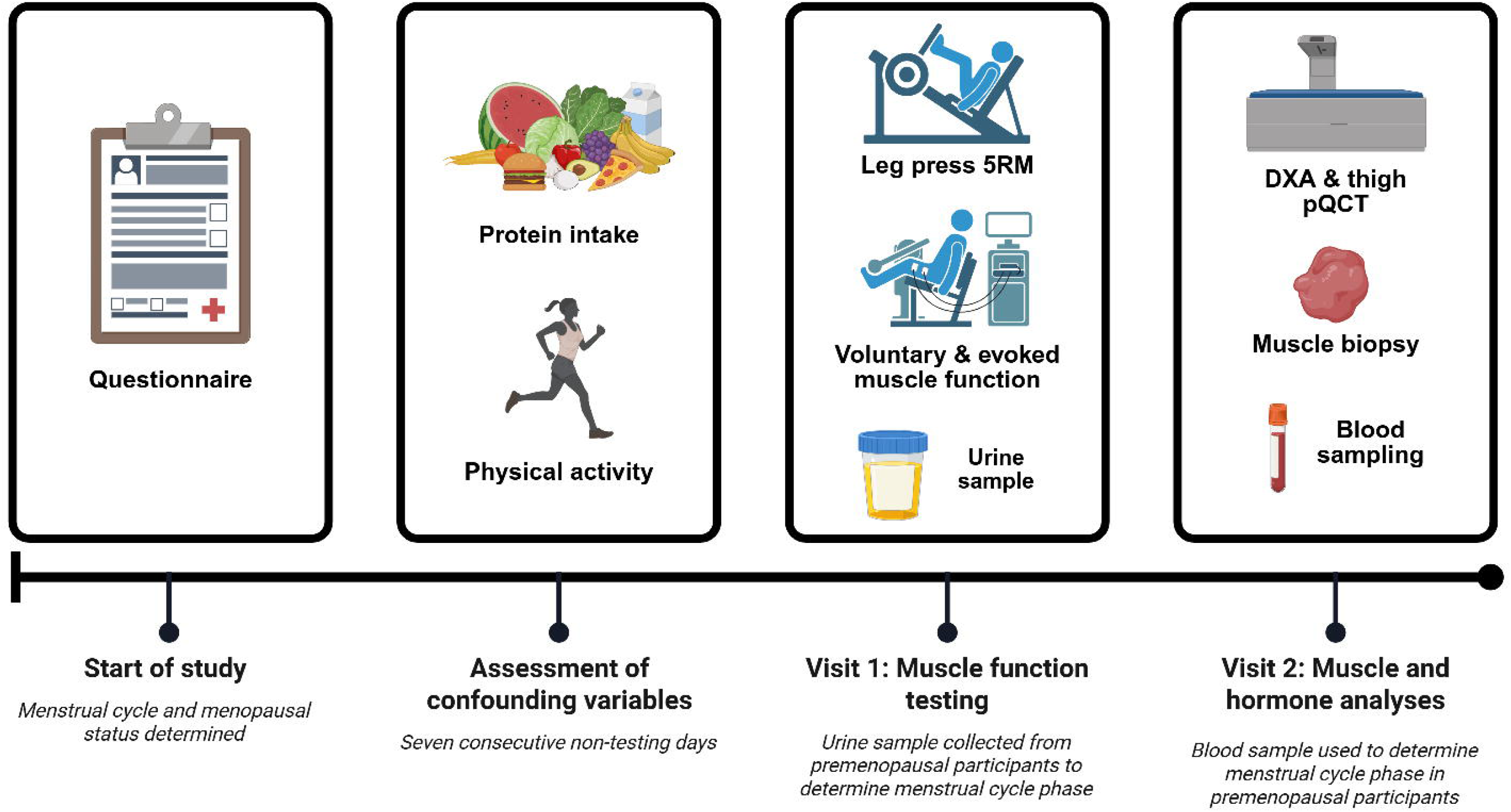
Study Outline. DXA: dual-energy x-ray absorptiometry; pQCT: peripheral quantitative computed tomography; 5RM: five-repetition maximum.

### Methodology

#### Muscle strength

Prior to visit one (Figure 1), participants were asked to abstain from vigorous exercise for 24 hours and to avoid caffeine on the day of the testing. At visit one, leg extensor strength was assessed via a leg press 5-repetition max (5-RM) test. Participants completed two warm-up sets of eight repetitions, followed by up to four test sets of five repetitions. After each test set, participants were asked to rate their perceived exertion from 6 to 20 on the Borg Scale (Borg, 1982). The resistance was increased until the participant was maximally exerted after five repetitions. Three participants did not complete the test. Additionally, some participants reached failure before completing 5 repetitions, so the weight and number of completed repetitions was recorded. From these values, a modified equation was used to calculate estimated 1-RM (e1RM, kg) (Wood *et al*., 2002):

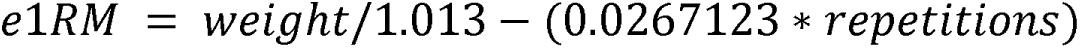

Isometric neuromuscular assessment of the quadriceps was conducted using an isokinetic dynamometer (Universal Pro Single Chair model 850-230, Biodex Medical Systems, United States), electrical stimulation, and surface electromyography, the precise and full methodology of which have been previously described in O’Bryan *et al*. (2025). Briefly, participants completed a warm up of submaximal and maximal isometric knee extensions and flexions. Following this, participants completed a ∼4s maximal isometric voluntary contraction of the knee extensors. An electrically evoked doublet (100 Hz) was applied to the femoral nerve at the plateau in voluntary torque, followed by three resting evoked twitch responses at 100 Hz, 10 Hz, and 1 Hz, ∼1.5s apart. This was repeated three times, each separated by two minutes of rest. The highest recorded maximal voluntary contraction (MVC, N.m), rate of torque development from the 1 Hz evoked twitch (N.m.s^-1^), and peak potentiated resting twitch torque at 100 Hz (PT_100Hz)_ and 10 Hz (PT_10Hz_) were included in this analysis.

#### Body composition

At visit two (mean interval = 6 days), participants arrived at the laboratory in the morning in a fasted state from midnight. They were given a standardised meal of pasta and tomato sauce to eat *ad libitum* the night before. Participants abstained from caffeine, alcohol, and vigorous physical activity in the 24 hours prior to the visit. Dual energy x-ray absorptiometry (DEXA; GE Lunar, Madison, WI) was used to assess body composition, including total body fat and lean mass (kg), and total body bone mineral content (BMC, kg). Participants lay in a supine anatomical position with the hands in a neutral position. A foam block was placed between the arms and trunk to separate the regions. The enCORE software (GE Healthcare, Chicago, IL) uses skeletal landmarks in the image to detect regions of interest, which were manually adjusted where appropriate. The leg region was determined by placing a border through the femoral neck and the arm region was defined by a border at the medial side of the humerus neck. The sum of leg and arm lean mass was used to calculate appendicular lean mass (ALM, kg), and ALM index was calculated by dividing ALM by height squared (m^2^).

XCT 3000 peripheral quantitative computed tomography (qPCT; Stratec Medizintechnik GmBH, Pforzheim, Germany) scans of the upper thigh were used to determine muscle cross-sectional area (CSA, cm^2^), subcutaneous and intramuscular fat area (cm^2^), and femur bone density (g.cm^-3^). Participants lay in a supine position with their measured leg secured in a footrest. Tibial length was used as an approximation of femur length, measured from the tibial plateau to the medial malleolus while the knee was flexed to 90° (Cervinka *et al*., 2018). The images were taken at 50% of the tibia length from the mid-condylar cleft towards the hip and analysed using ImageJ software (version 2.0.0; National Institutes of Health, Bethesda, MD, USA). Movement artifacts on the scan were scored from one (none) to five (extreme) based on criteria from Blew *et al*. (2014). Scans with a score ≥ four were excluded from the analysis (n = 11). Thigh muscle percentage (%) was calculated by dividing muscle CSA (cm^2^) by total thigh area (cm^2^) and multiplying by 100. The effective radiation dose received by participants undergoing the DXA and pQCT scans was approximately 0.04 mSv.

#### Blood and muscle sampling

In the same visit, 5mL of blood was taken from the antecubital vein, centrifuged at 4°C to isolate the plasma, and stored at -80°C until required for analysis. Hormone analysis was completed at Monash Health Pathology laboratory (Victoria, Australia). Total E2 (pmol/L) and testosterone (TT, nmol/L) were measured using high-performance gas chromatography mass spectrometry (Triple Quad 5500, Sciex, Framingham, MA). Progesterone (nmol/L), luteinising hormone (LH, IU/L), follicle-stimulating hormone (FSH, IU/L), and sex hormone-binding globulin (SHBG, nmol/L) were measured using a sequential two-step immunoenzymatic sandwich assay (UniCel DxI 800 Access Immunoassay System, Beckman Coulter, New South Wales, Australia). Free E2 index (FEI) was calculated by dividing E2 (nmol/L) by SHBG (nmol/L) and multiplying by 100. Free androgen index (FAI) was calculated by dividing total testosterone (nmol/L) by SHBG (nmol/L) and multiplying by 100. The testosterone to E2 (T/E2) ratio was calculated by dividing total testosterone (nmol/L) by total E2 (nmol/L).

A muscle biopsy was then taken from the *vastus lateralis* using the percutaneous needle technique modified to include suction (Evans *et al*., 1982). First, the skin was sterilised and anaesthetised with 1% lidocaine. Following this, an incision was made through the skin and fascia. The muscle sample was removed, immediately snap frozen in liquid isopentane, and stored at -80°C until needed for analysis.

#### Control of confounding variables

To minimise fluctuations in sex hormones as a potential confounding variable amongst premenopausal females, we aimed to perform tissue/blood sampling and muscle strength testing within the first seven days of their self-reported menstrual cycle, where the ratio of oestrogen to progesterone is the lowest and most stable, and therefore can be considered to be representative of ‘baseline’ concentrations (Elliott-Sale *et al*., 2021). This phase is also easiest to identify due to the presence of obvious symptoms (e.g. menstrual bleeding or discharge). To account for the cases where this was not possible due to unpredictable cycle length and/or masking of usual menstrual symptoms by hormonal contraception, premenopausal participants collected an additional urine sample on the morning of visit one, which was used to quantify E2 (Abcam, Cambridge, United Kingdom) and progesterone (Invitrogen, Waltham, MA) with enzyme linked immunosorbent assays (ELISA) as described in O’Bryan *et al*. (2025). Plasma and urinary hormone levels were then used in addition to a cycle calendar (e.g. last known period, day of cycle) to determine menstrual cycle phase for each visit, using a modified version of Elliot-Sale *et al*. (2021), as displayed in Table 1.

**Table 1.**
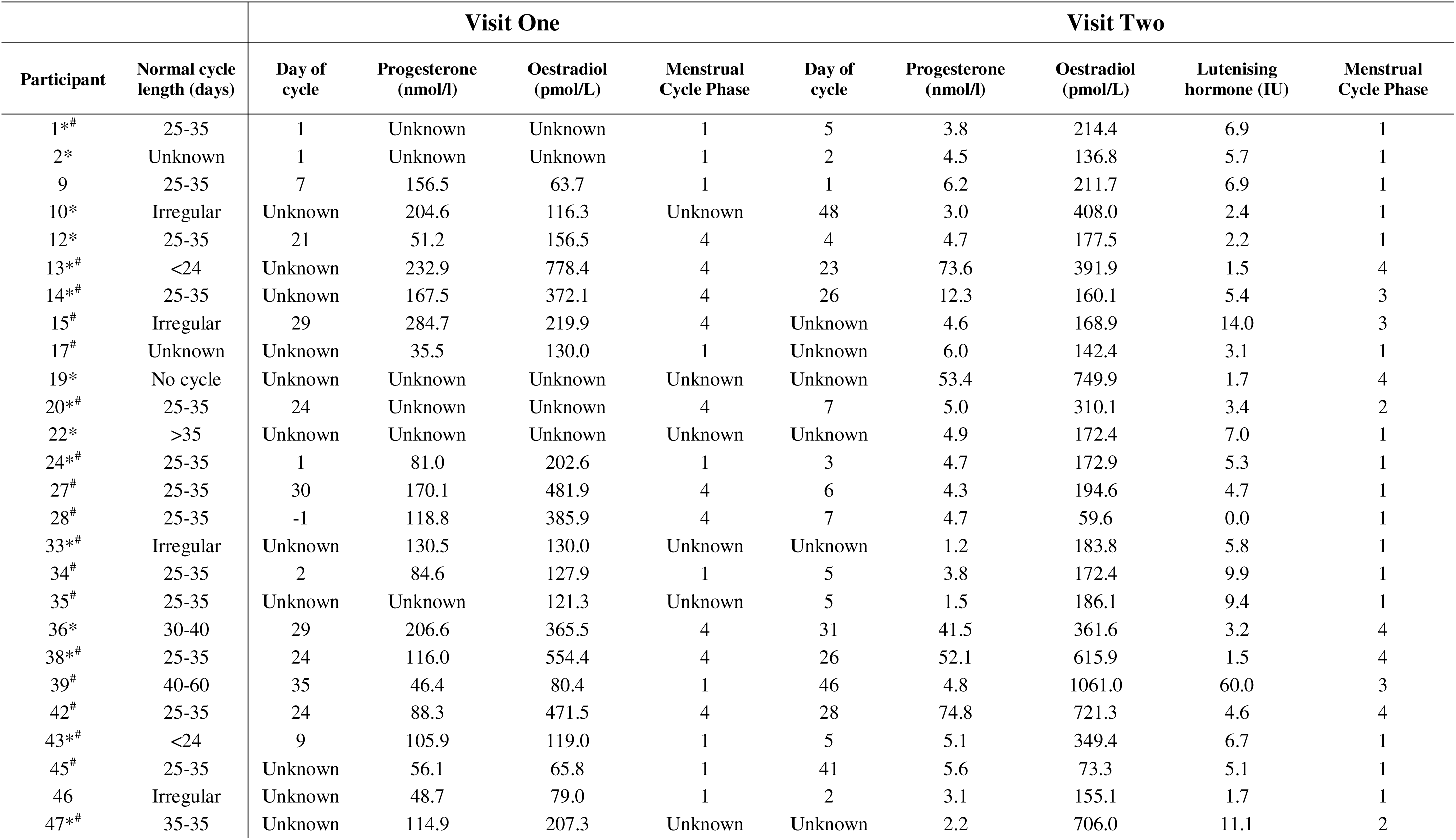
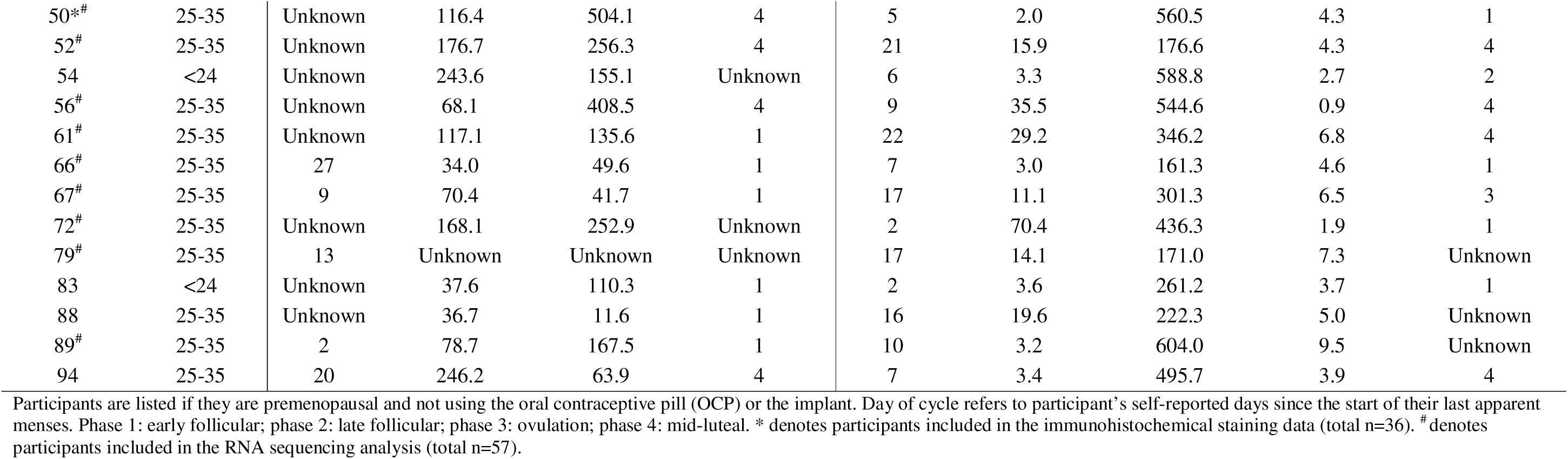
Menstrual cycle phase at each study visit.

Additional confounding variables including MVPA (moderate-to-vigorous physical activity) and protein intake were measured over a 7-day period. Participants were asked to record their physical activity with an ActiCal accelerometer watch (Respironics Inc, Murraysville, PA). During the hours in which participants were awake (determined by a sleep diary) the time spent in moderate and vigorous physical activity (>1,535 counts per minute; MVPA) per day was calculated and averaged across all valid days (Colley & Tremblay, 2011). Days were considered valid if there was ≥10 hours of data available, and participants required at least 4 valid days of data to be included in the physical activity analysis. During the same period, participants were asked to record all food and drink intake via the Easy Diet Diary app (Xyris Software, Brisbane, Australia). Average protein intake (g/kg body weight/day) was calculated from participants with at least 4 full days of data. Twelve participants did not meet the MVPA data criteria, and 15 did not meet the food diary criteria.

### Skeletal muscle analysis

#### Immunohistochemical staining

In a subset of 36 participants (n = 6 per decade of age randomly selected determined by a power calculation based on Léger *et al*., 2008), *vastus lateralis* muscle samples were immunohistochemically stained to determine muscle fibre size and type. Briefly, samples were cut into 10 µm sections using a cryostat at -20°C and stored at -80°C until required. Upon use, sections were thawed at room temperature for 10 minutes and incubated for one hour in blocking solution (10% goat serum in 1 x phosphate buffered saline (PBS)). Sections were then incubated for one hour with a primary antibody cocktail in blocking solution: BA-F8 (type I fibres, 1:20, Developmental Studies Hybridoma Bank (DSHB), University of Iowa, IA), SC-71 (type IIa fibres, 1:50, DSHB, University of Iowa, IA), and α-laminin (cell membrane, 1:100, Merck, Burlington, MA). After washing with PBS, sections were incubated for one hour in the dark with a solution of secondary antibodies in PBS: goat α-mouse IgG2b Alexa Fluor 647 (1:500, Life Technologies, Carlsbad, CA), goat α-mouse IgG1 Alexa Fluor 488 (1:500, Life Technologies, Carlsbad, CA), and goat α-rabbit IgG Alexa Fluor 405 (1:500, Life Technologies, Carlsbad, CA). The sections were washed, mounted, and imaged using the Eclipse Ti2 confocal microscope (Nikon, Tokyo, Japan) at 10x magnification. Regions of intact, cross-sectional fibres were selected to create regions of interest using Fiji (ImageJ) (Schindelin *et al*., 2012) and analysed using the SMASH (semi-automated muscle analysis using segmentation of histology) application on MATLAB (R2023b, The MathWorks Inc., Natick, MA) to quantify fibre CSA and relative the proportions of each fibre type (I, IIa, I/IIa, and IIx) by fibre number and area. Hybrid I/IIa fibres were defined as fibres that were positively stained for both BA-F8 and SC-71. Unstained fibres were considered type IIx but their occurrence was very rare (<0.5%), so they were not included in the analysis. The average number of fibres per section was 331 ± 177.

To quantify satellite cells, a second batch of muscle sections were thawed and fixed in ice cold acetone for three minutes. Following a wash with PBS, endogenous peroxidases were blocked with 3% H_2_O_2_ in PBS for seven minutes at room temperature and washed again. The muscle sections were incubated for one hour in 2.5% normal horse serum (NHS), followed by an overnight incubation with α-laminin (1:100, Merck, Burlington, MA) and α-Pax7 (1:100, DHSB, University of Iowa, IA) in 2.5% NHS. After washing, the sections were incubated for one hour in the dark with goat α-rabbit IgG Alexa Fluor 647 (1:250, Invitrogen, Waltham, MA) and goat α-mouse IgG biotin-SP-conjugated (1:1000, Jackson ImmunoResearch Labs, Westgrove, PA) in PBS. The sections were subsequently incubated in PBS with streptavidin-horseradish peroxidase conjugate (one hour, 1:100, Invitrogen, Waltham, MA), Alexa Flour 488 tyramide (20 minutes, 1:200, Invitrogen, Waltham, MA), and DAPI (4’,6-diamidino-2-phenylindole, 10 minutes, 1:10,000, Invitrogen, Waltham, MA), each separated by a wash, and finally mounted. The sections were imaged at 10x magnification using the tiles and stitching functions on an AxioImager M2 upright microscope (Zeiss Zen 3.1, Oberkochen, Germany). Satellite cells were defined as Pax7+/DAPI+ cells located on the laminin border and were normalised to fibre number and total section area.

To quantify intermyofibrillar adipose tissue (IMAT) and fibrosis, a third batch of *vastus lateralis* muscle sections (10 µm thick) were thawed and fixed in 4% paraformaldehyde for seven minutes. Following a wash in PBS, the sections were blocked in 2.5% normal horse serum for one hour, followed by an overnight incubation with α-perilipin-1 (1:100, Abcam, Cambridge, UK) in 2.5% normal horse serum. After a wash, the sections were incubated in the dark for an hour with goat α-rabbit Alexa Fluor 555 (1:250, Invitrogen, Waltham, MA) and wheat germ agglutinin (WGA) Alexa Fluor 647 (1:50, Thermo Fisher Scientific, Waltham, MA) in PBS, washed again, and incubated for 30 minutes with boron dipyrromethane (BODIPY, 1 mg/mL ethanol, 1:50, Life Technologies, Carlsbad, CA) in PBS. Finally, the sections were washed and mounted with Vectashield Plus mounting medium with DAPI (Vector Laboratories, Newark, CA) and imaged at 10x magnification using the tiles and stitching functions on an AxioImager M2 upright microscope (Zeiss Zen 3.1, Oberkochen, Germany). Image analysis was performed in a blinded manner. WGA was quantified using the threshold feature in ImageJ (Schindelin *et al*., 2012), and the area occupied by WGA was expressed as a percentage relative to the total muscle area per our previous publication (Noehren *et al*., 2021). This value is reported as fibrosis percentage. BODIPY was quantified using custom macro scripts in Fiji. Briefly, image files were uploaded to Fiji and separated into BODIPY and WGA channels. A myofibre mask was generated by taking a threshold of the WGA channel. This mask was used to discern BODIPY signal that resided inside and outside the myofibres. Only BODIPY signal located outside myofibres was considered for the analysis of IMAT. After setting a separate threshold for the BODIPY channel which filtered out background noise, the number of pixels with positive BODIPY signal that resided within the muscle cross-section, but not inside the myofibres, was calculated and divided by the total pixel area of the muscle cross-section. This value represents the percentage of BODIPY positive area and is reported as IMAT percentage.

#### RNA sequencing

Forty-eight participants (n = 8 per age group) were randomly selected for muscle RNA sequencing using stratified randomisation with random sorting, and were added to 10 participants from this same cohort (aged between 19 and 37 years), who had already undergone muscle RNA sequencing for a previous study (Lamon *et al*., 2024), resulting in a total of 58 samples. RNA was extracted from 10mg of snap frozen *vastus lateralis* tissue, using the AllPrep DNA/RNA/miRNA Universal kit (Qiagen, Hilden, Germany). cDNA libraries were prepared using the Ribo-Zero Gold kit and the Illumina TruSeq Stranded Total RNA protocol (Illumina, San Diego, CA). RNAseq libraries were prepared using the Illumina TruSeq Stranded Total RNA with Ribo-Zero Gold protocol and sequenced with 150-bp paired-end reads on the Illumina Novaseq6000 (Macrogen Oceania Platform). Reads underwent quality check with FastQC (v0.11.9; Babraham Institute, Cambridge, UK) and Kallisto (v0.46.1) was used to map reads to the human reference genome (*HomoSapien GRCh38)* and to generate transcript counts. One sample was excluded from analysis as it did not pass quality control due to poor alignment. The counts matrices from both batches of samples were merged into one matrix. PCA plots made using the RNAseqQC package (v0.2.1) (DeLuca *et al*., 2012) highlighted significant batch effects, which were subsequently removed by adding batch number as a covariate in all differential expression (DE) analyses and confirmed with a PCA plot. Genes with an average of 10 reads per million (RPM) or less, across all samples, were removed from further analysis. 16,928 Ensemble gene IDs were used in downstream analysis.

### Statistical analysis

#### Regression analysis

All statistical analysis was performed using Rstudio 4.4.2 (R Core Team, 2021). Firstly, linear regressions were used to determine potential associations between hormonal contraception, hormone replacement therapy use or menstrual cycle phase on body composition, muscle morphology, or muscle strength outcomes.

All body composition, muscle morphology and muscle strength outcomes were then plotted against age and assessed for linearity by evaluating heteroscedasticity of residuals and the Akaike information criterion (AIC) with a linear or polynomial regression model, after adjusting for MVPA and dietary protein intake. These covariates were selected as they can both significantly impact muscle metabolism (McGlory *et al*., 2019). The model with the best fit (i.e., lower heteroscedasticity and AIC score) was selected. Linear models were selected for all outcome variables. Normality of residuals was confirmed by plotting a histogram of studentised residuals. Homoscedasticity was assessed with a plot of fitted versus predicted residuals. Missing covariate data (MVPA and protein intake) were imputed using the kNN function in the VIM package (Kowarik & Templ, 2016). Multicollinearity of covariates was assessed with variation inflation factors (VIF); a value of 3 was used as a threshold for excluding covariates from the model.

Multiple linear regression was also used to determine the associations between circulating sex hormones and all outcome measures. Body composition and muscle morphology models were adjusted for age, MVPA, and protein intake. Muscle function models were adjusted for age, MVPA, protein intake and thigh subcutaneous fat, and were assessed in postmenopausal females only, as muscle function testing occurred on a different day to hormone quantification, so we could not rule out confounding effects of menstrual cycle hormone fluctuations. Some heteroscedasticity was observed, so robust standard errors were used. Z-scores were calculated for all independent, dependent, and confounding variables and used in the sex hormone regression models to allow comparison between effect sizes. A z-score of 0 indicates a value equal to the mean, and z-score of 1 indicates a value equal to the standard deviation. A ‘weak’ association was defined as a standardised coefficient <0.20, a ‘moderate’ association was defined as a standardised coefficient between 0.20 to 0.50, and a ‘strong’ association was defined as a standardised coefficient of >0.50.

#### Transcriptomics analysis

The DESeq2 package (v1.46.0) was used to perform normalisation and differential expression analysis (Love et al., 2014). For data visualization and dimensionality reduction, the variance stabilizing transformation (VST) was applied to the filtered count matrix to reduce heteroscedasticity prior to principal component analysis (PCA). For differential expression analysis, DESeq2’s internal normalisation via size factor estimation was used. Benjamini–Hochberg adjusted p-values <0.05 and absolute log2 fold change >1 were considered significant. Two models were used: 1) to identify genes with age-associated expression changes using the design formula ∼ batch + age, and 2) to identify associations between gene expression and standardized circulating sex hormone concentrations using ∼ batch + hormone. Both models treat the covariate as continuous and assume a linear relationship on the log scale of the generalised linear model. Pathway enrichment was conducted on all genes that were significantly associated with age using the gseGO function from the clusterProfiler package (v4.14.6).

The Likelihood Ratio Test (LRT) in the DESeq2 package was used to identify how genes are differentially expressed between individual age groups (18-29, 30-39, 40-49, 50-59, 60-69, and 70-80 years), and those with similar patterns of expression were clustered using the degPatterns function from the DEGreport package (v1.42.0). Pathway enrichment analysis was conducted to determine the pathways represented in each cluster. The enricher function from the clusterProfiler package (v4.14.6) was used with the Reactome gene sets from *Homo sapiens* using the msigdbr package (v7.5.1), with a minimum gene set size of 5 and an adjusted *p*-value cutoff of FDR < 0.05.

Enrichment of transcription factor binding sites (TFBS) was conducted with the UniBind enrichment analysis tool, which uses the runLOLA function of the LOLA package (https://unibind.uio.no/) (Puig *et al*., 2021). The analysis type used was enrichment with a background, with a p-value threshold of <0.01 and inclusion of transcription factors (TFs) from all tissues. DEseq2 was first used to determine differentially expressed genes (DEGs) between young (18-30 years) and old (60-80 years) participants (expression ∼ batch + age group), and between pre- and post-menopausal participants (expression ∼ batch + menopausal status). Promotor regions of genes that were significantly upregulated compared to the contrast group underwent enrichment of TFBS against a background of the promotor regions of all expressed genes.

Due to the unique multinucleated nature of skeletal muscle cells, there is currently no method allowing quantitative deconvolution of bulk RNA-Seq data in skeletal muscle (Conning-Rowland et al., 2025; Voisin et al., 2024), however, MuSiC (Multi-subject Single Cell deconvolution) and other algorithms allow to draw robust semi-quantitative comparisons between groups of samples, or between different muscle sample populations (Conning-Rowland et al., 2025). Deconvolution of the bulk skeletal muscle transcriptome data was performed using the MuSiC package in R (Wang *et al*., 2019). As a reference, publicly available single-cell RNA-seq data were used from the Tabula Sapiens atlas, specifically the skeletal muscle dataset (“TS_Muscle.h5ad” available via FigShare). Preprocessing of the single-cell data was conducted using Scanpy in Python (v3.12). Cells with fewer than 200 expressed genes and genes expressed in fewer than 3 cells were filtered out to remove low-quality data. Raw (unnormalised) read counts from both the bulk and single-cell datasets were used as input to the music_prop() function, with free_annotation as the clustering variable and donor as the subject identifier. Cell type proportions were estimated and reported as decimals, representing relative abundance per sample. For ease of interpretation, the cell types were manually grouped into six biologically meaningful categories: immune, endothelial, muscle, stromal, epithelial, and erythrocyte. The RNA content of each muscle cell type was normalised to the total muscle RNA content to account for the multinuclearity of skeletal muscle fibres. The proportions of each cell type were then normalised to the mean of the 18–29-year-old age group. One-way analysis of variance (ANOVA) was used to determine a significant difference across all age groups, with a *p*-value threshold of < 0.05.

## Results

### Participant characteristics

Ninety-six participants completed the study, with every decade of age between 18-80 represented in the cohort. Forty-seven participants were premenopausal, four were perimenopausal, and 45 were postmenopausal. The early-follicular phase could not be unequivocally confirmed in 23 and 19 participants at visit one and two, respectively (Table 1). All participant characteristics are displayed in Table 2. The two sub-sets of participants that underwent muscle fibre histological analysis and RNA sequencing are characterised in Table A1-A2. There was no association between menstrual cycle phase on any measure of muscle strength (*p*>0.05, Table 3). There was also no association between hormonal contraception (oral contraceptive pill, intrauterine device, or implant) or hormone replacement therapy and any measure of body composition, or muscle strength in pre- or post-menopausal participants, respectively (*p*>0.05). In premenopausal participants, OCP and IUD use was significantly associated with greater type I muscle fibre CSA (*p*=0.044 and *p*=0.014, respectively; Table 3). No other muscle morphology measures were significantly associated with hormonal contraceptive use.

**Table 2.** Participant characteristics.

| Variable | Overall<br>n = 96 | 18-29<br>n = 21 | 30-39<br>n = 13 | 40-49<br>n = 15 | 50-59<br>n = 16 | 60-69<br>n = 15 | 70-80<br>n = 16 |
| --- | --- | --- | --- | --- | --- | --- | --- |
| **Body composition** |  |  |  |  |  |  |  |
| Weight (kg) | 66.1 (12.2) | 64.6 (13.6) | 66.4 (13.6) | 68.0 (12.2) | 65.0 (12.5) | 68.5 (9.6) | 64.7 (12.5) |
| Height (m) | 1.64 (0.07) | 1.64 (0.08) | 1.64 (0.8) | 1.66 (0.07) | 1.65 (0.07) | 1.65 (0.06) | 1.60 (0.5) |
| BMI (kg.m<sup>-2</sup>) | 24.5 (4.3) | 23.9 (4.6) | 24.8 (4.4) | 24.7 (4.0) | 23.8 (3.9) | 25.2 (4.8) | 25.2 (4.6) |
| Body fat (%) | 33.5 (8.4) | 30.9 (6.9) | 33.9 (9.5) | 32.9 (8.6) | 32.1 (8.4) | 36 (8.9) | 36 (8.6) |
| Total body fat mass (kg) | 21.8 (9.1) | 19.3 (8) | 22.5 (10.4) | 21.8 (8.7) | 20.6 (9.8) | 24.3 (9.3) | 23.3 (9.3) |
| Total body lean mass (kg) | 41.4 (5.3) | 41.9 (7.4) | 41.1 (4.6) | 43.3 (5.7) | 41.6 (4.6) | 41.5 (3.9) | 38.8 (3.7) |
| ALM (kg) | 18.3 (2.9) | 19.1 (3.9) | 18.3 (2.5) | 19.1 (2.8) | 18.3 (2.5) | 18.1 (2.0) | 16.7 (2.3) |
| ALM index (kg.m<sup>-2</sup>) | 6.8 (0.9) | 7.1 (1.2) | 6.8 (0.8) | 6.9 (0.8) | 6.7 (0.6) | 6.7 (0.6) | 6.5 (0.9) |
| Bone mineral content (kg) | 2.3 (0.4) | 2.4 (0.4) | 2.4 (0.3) | 2.5 (0.3) | 2.3 (0.3) | 2.2 (0.3) | 2.1 (0.3) |
| **Muscle size and composition** |  |  |  |  |  |  |  |
| Thigh muscle CSA (cm<sup>2</sup>) | 92.2 (17.8) | 102.6 (19.4) | 99.4 (16.1) | 99.8 (13.3) | 85.8 (13.1) | 88.2 (13.1) | 74 (12.9) |
| Intramuscular fat (cm<sup>2</sup>) | 3.7 (2.5) | 3.2 (2.9) | 2.7 (1.6) | 3.3 (2.3) | 3.5 (1.4) | 5.0 (3.0) | 4.6 (2.4) |
| Subcutaneous fat (cm<sup>2</sup>) | 69.7 (30.8) | 70.2 (34.3) | 68.6 (28.1) | 71.1 (32.2) | 59.7 (12.4) | 72.1 (36.4) | 78.4 (37.1) |
| Relative thigh muscle (%) | 53.7 (16.7) | 55.0 (8.9) | 54.8 (8.7) | 54.1 (9.5) | 61.6 (33.9) | 50.3 (10.2) | 44.6 (7.4) |
| Femur bone density (g.cm<sup>-3</sup>) | 781.5 (69.3) | 812.2 (63.6) | 803.0 (63.2) | 813.9 (43.7) | 785.8 (42.4) | 750.5 (90.3) | 713.1 (38.3) |
| **Muscle strength** |  |  |  |  |  |  |  |
| Leg press e1RM (kg) | 83.9 (25.5) | 101.7 (33.7) | 90 (24.3) | 90.2 (20.7) | 79.8 (14.6) | 72.1 (12.3) | 62.4 (12.3) |
| Specific leg press e1RM |  |  |  |  |  |  |  |
| *Normalised to leg lean mass (kg.kg<sup>-1</sup>)* | 6 (1.5) | 6.9 (1.9) | 6.4 (1.3) | 6.1 (0.9) | 5.8 (1.2) | 5.3 (0.9) | 4.9 (1) |
| *Normalised to thigh muscle CSA (kg.cm<sup>-2</sup>)* | 0.9 (0.2) | 1 (0.2) | 0.9 (0.2) | 1 (0.2) | 0.9 (0.2) | 0.8 (0.2) | 0.8 (0.1) |
| Maximal voluntary contraction (N.m) | 163.83 (53.5) | 156.1 (54.5) | 165.7 (33.5) | 138.3 (26.2) | 139.6 (39.2) | 109.6 (22.8) | 146.2 (44.0) |
| Rate of torque development (N.m.s<sup>-1</sup>) | 1,400.4 (345.0) | 1,450.6 (182.0) | 1,431.8 (226.0) | 1,228.7 (220.2) | 1,208.2 (177.2) | 1,065.4 (220.5) | 1,306.8 (271.9) |
| PT<sub>100Hz</sub> (N.m) | 65.0 (13.7) | 67.8 (8.6) | 66.8 (9.6) | 57.2 (7.7) | 60.8 (10.5) | 48.9 (8.6) | 61.5 (11.7) |
| PT<sub>10Hz</sub> (N.m) | 62.3 (12.8) | 69.8 (13.1) | 64.7 (11.5) | 57.1 (9.0) | 60.3 (11.4) | 47.7 (10.6) | 60.7 (12.9) |
| PT<sub>100Hz:10Hz</sub> ratio | 1.05 (0.12) | 0.98 (0.08) | 1.04 (0.11) | 1.01 (0.05) | 1.01 (0.06) | 1.04 (0.09) | 1.02 (0.09) |
| **Circulating sex hormones** |  |  |  |  |  |  |  |
| Total E2 (pmol/L) | 192.2 (203.5) | 195 (156) | 355.6 (283.3) | 366.6 (250.4) | 125 (130.0) | 74.7 (18.8) | 69.6 (39.7) |
| Free E2 index (FEI) | 0.49 (0.81) | 0.64 (1.24) | 1.04 (1.01) | 0.71 (0.68) | 0.32 (0.45) | 0.16 (0.08) | 0.12 (0.09) |
| Total progesterone (nmol/L) | 9.2 (16.3) | 10.7 (17.6) | 15.3 (21.2) | 20.5 (25.8) | 3.3 (2.2) | 3.5 (2) | 2.7 (1.4) |
| Total testosterone (nmol/L) | 0.81 (0.39) | 0.96 (0.38) | 0.93 (0.44) | 0.73 (0.21) | 0.67 (0.33) | 0.72 (0.36) | 0.84 (0.51) |
| Free androgen index (FAI) | 1.8 (1.5) | 2.4 (2.2) | 2.7 (1.7) | 1.3 (0.8) | 1.5 (0.9) | 1.4 (0.8) | 1.4 (1.0) |
| TT/E2 ratio | 8.1 (7.4) | 6.7 (3.4) | 3.8 (2.6) | 3.8 (3.3) | 8.7 (6.5) | 11.1 (10.5) | 14 (9.5) |
| **Reproductive health characteristics** |  |  |  |  |  |  |  |
| Menopausal status |  |  |  |  |  |  |  |
| *Premenopausal* | 47 / 96 (49%) | 21 / 21 (100%) | 13 / 13 (100%) | 12 / 15 (80%) | 1 / 16 (6%) | - | - |
| *Perimenopausal* | 4 / 96 (4%) | - | - | 2 / 15 (13%) | 2 / 16 (13%) | - | - |
| *Postmenopausal* | 45 / 96 (47%) | - | - | 1 / 15 (7%) | 13 / 16 (81%) | 15 / 15 (100%) | 16 / 16 (100%) |
| Hormonal contraception |  |  |  |  |  |  |  |
| *None* | 31 / 96 (32%) | 10 / 21 (48%) | 8 / 13 (62%) | 10 / 15 (67%) | 3 / 16 (19%) | - | - |
| *OCP* | 8 / 96 (8%) | 5 / 21 (24%) | 1 / 13 (8%) | 2 / 15 (13%) | - | - | - |
| *IUD* | 11 / 96 (11%) | 5 / 21 (24%) | 4 / 13 (31%) | 2 / 15 (13%) | - | - | - |
| *Implant* | 1 / 96 (1%) | 1 / 21 (5%) | - | - | - | - | - |
| *N/A (postmenopausal)* | 45 / 96 (47%) | - | - | 1 / 15 (7%) | 13 / 16 (81%) | 15 / 15 (100%) | 16 / 16 (100%) |
| Hormonal replacement therapy |  |  |  |  |  |  |  |
| *Currently using* | 9 / 96 (9%) | - | - | 2 / 15 (13%) | 2 / 16 (13%) | 3 / 15 (20%) | 2 / 16 (13%) |
| *Not currently using* | 40 / 96 (42%) | - | - | 1 / 15 (7%) | 13 / 16 (81%) | 12 / 15 (80%) | 14 / 16 (88%) |
| *N/A (premenopausal)* | 47 / 96 (49%) | 21 / 21 (100%) | 13 / 13 (100%) | 12 / 15 (80%) | 1 / 16 (6%) | - | - |
| **Lifestyle factors** |  |  |  |  |  |  |  |
| Dietary protein intake (g.kg<sup>-1</sup>.day<sup>-1</sup>) | 1.30 (0.42) | 1.31 (0.52) | 1.56 (0.44) | 1.28 (0.42) | 1.45 (0.34) | 1.16 (0.31) | 1.17 (0.43) |
| MVPA (mins.day<sup>-1</sup>) | 36.8 (29.6) | 42.4 (23.9) | 39 (24.1) | 34.9 (42.5) | 43.7 (20.3) | 39.2 (34.5) | 20.4 (27.1) |

Presented as mean (SD). ALM: appendicular lean mass; CSA: cross-sectional area; e1RM: estimated one-repetition maximum; E2: oestradiol; IUD: intrauterine device; MVPA: moderate-to-vigorous physical activity; OCP: oral contraceptive pill; TT: total testosterone.

**Table 3.**
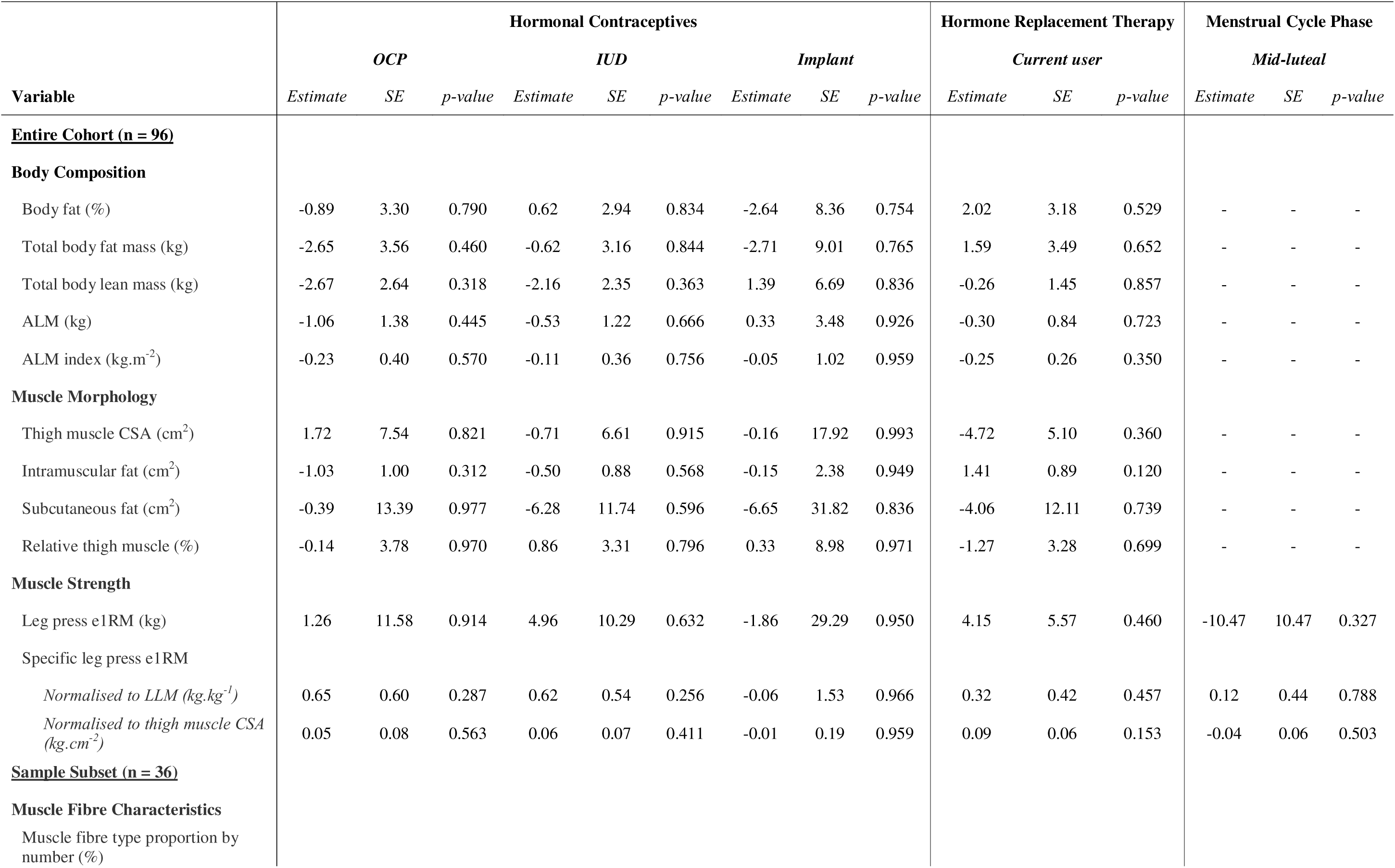
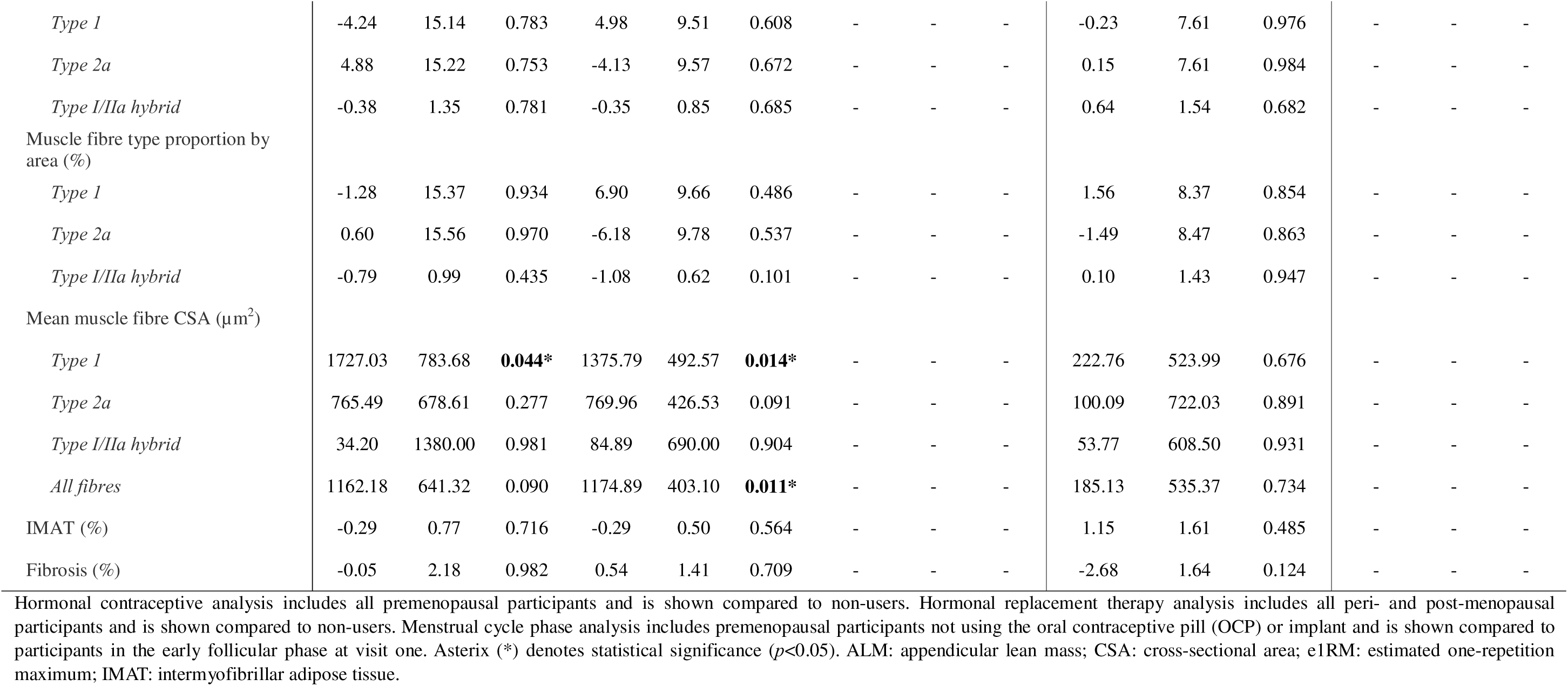
Associations between use of hormonal contraceptives, hormone replacement therapy and menstrual cycle phase on muscle outcomes.

### Female body composition and muscle strength across the lifespan

Linear associations between age and body composition, muscle morphology, and muscle strength are displayed in Fig. 2. Results from the unadjusted analysis are displayed in Table 3A. After adjusting for MVPA and protein intake, there was a significant negative association for age with ALM (*p*=0.022; Fig. 2B), thigh muscle CSA (*p*<0.001; Fig. 2D), thigh muscle percentage (*p*<0.007; Fig. 2G), e1RM (*p*<0.001; Fig. 2H), and specific e1RM normalised to both leg lean mass (*p*<0.001; Fig. 2I) and thigh muscle CSA (*p*<0.012; Fig. 2J), while age was positively associated with thigh intramuscular fat (*p*<0.014; Fig. 2F). Age-related changes in quadriceps MVC, RTD, PT_100Hz_ and PT_10Hz_ for this cohort were non-linear and have been reported elsewhere (O’Bryan *et al*., 2025).

**Figure 2.**
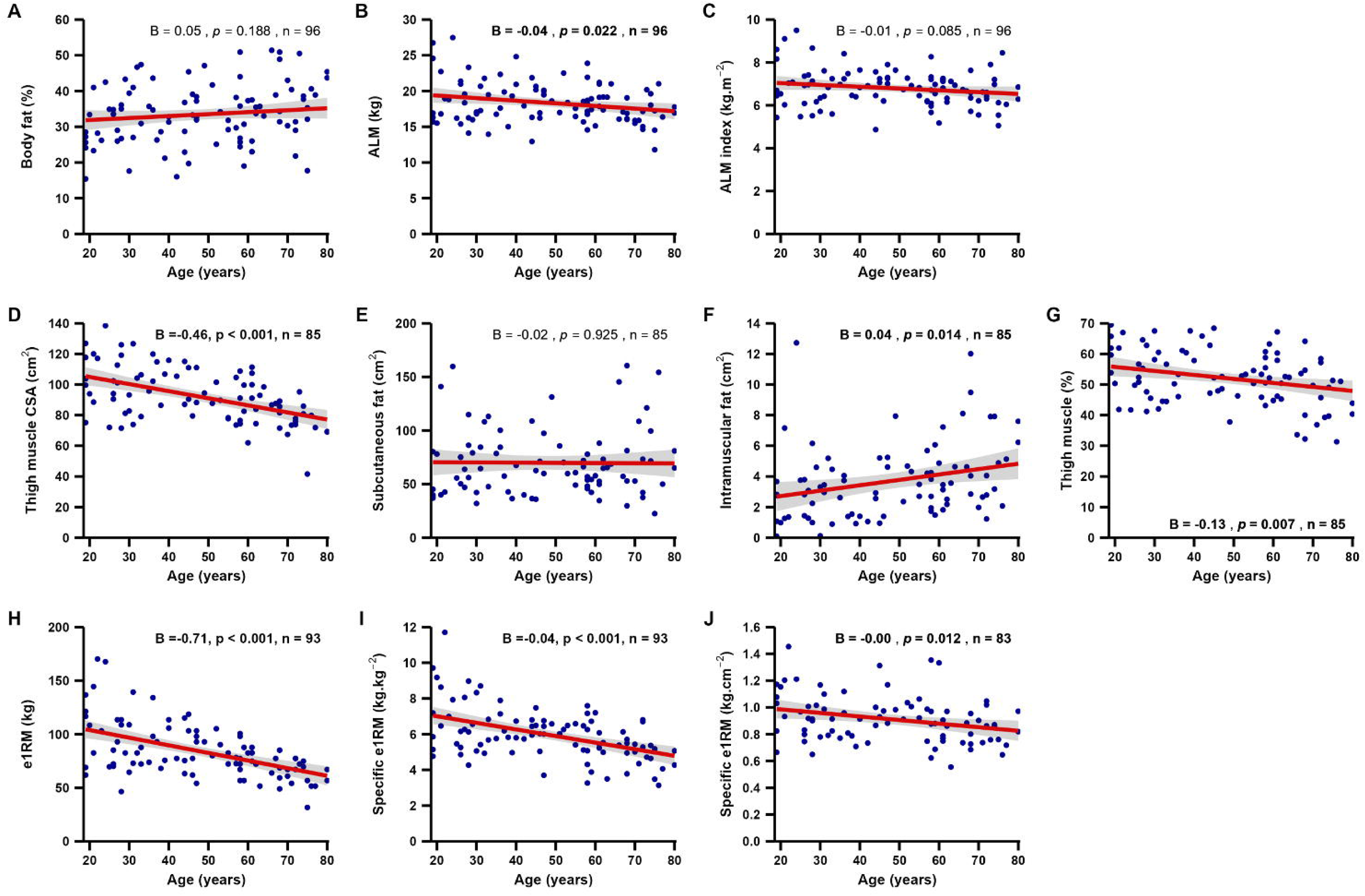
Age and female body composition, muscle morphology, and muscle strength. A-J) Linear associations between age and body composition and muscle strength. Red line represents B (coefficient) after adjusting for MVPA (moderate-to-vigorous physical activity) and protein intake. Specific e1RM normalised to leg lean mass (I) and thigh muscle cross-sectional area (J). Statistical significance accepted as *p*<0.05. ALM: appendicular lean mass; CSA: cross-sectional area; e1RM: estimated one-repetition maximum.

### Female muscle morphology across the lifespan

Linear associations between age and *vastus lateralis* muscle fibre characteristics in a random subset of 36 participants stratified by decade of age are displayed in Fig. 3. Unadjusted models are shown in Table A3. After adjusting for MVPA and dietary protein intake, there was a positive linear association between age and the proportion of hybrid fibres by area (*p*=0.038; Fig. 3F) and fibrosis (p=0.023; Fig. 3M) and there was a negative linear association between age and type I fibre CSA (*p*<0.001; Fig. 3G). None of the other muscle morphology outcomes were significantly associated with age (*p*>0.05).

**Figure 3.**
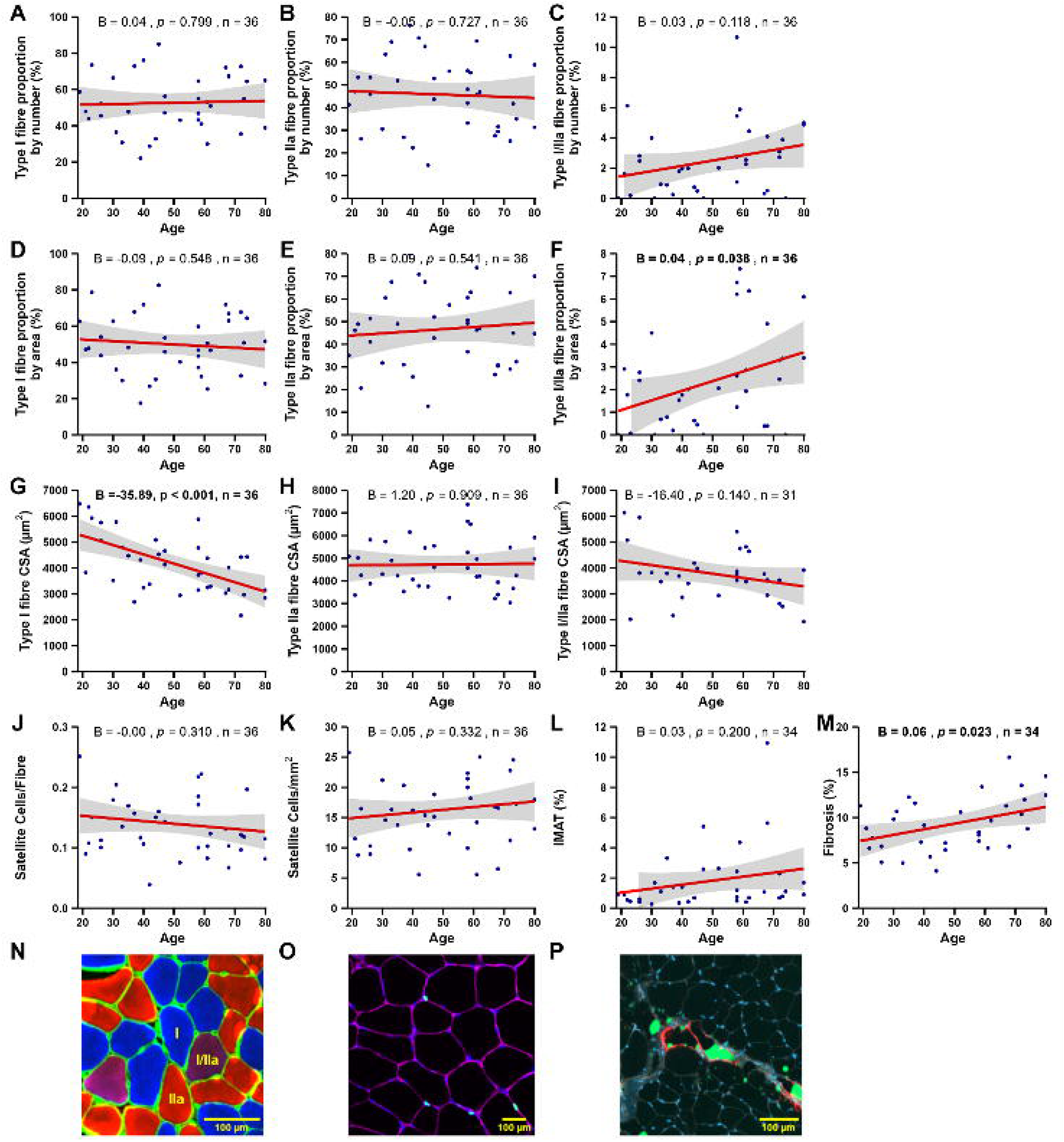
Age and female muscle morphology. A-M) Linear associations between age and muscle fibre characteristics. Red line represents B (coefficient) after adjusting for MVPA (moderate-to-vigorous physical activity) and protein intake. Statistical significance accepted as *p*<0.05. N) Representative image of muscle fibre type immunohistochemical staining, showing type I (blue), type IIa (red), and hybrid I/IIa (purple) fibres. O) Representative image of satellite cell immunohistochemical staining, showing Pax7^+^ cells (turquoise), laminin (purple), and DAPI (blue). P) Representative image of BODIPY (green), perilipin (red), wheat germ agglutinin (WGA, grey) and DAPI (blue) staining. All images taken at 10 x magnification. All models were adjusted for age, moderate-vigorous physical activity (MVPA), and protein intake.

### Associations between circulating sex hormones and female muscle

Associations between circulating sex hormones and body composition and muscle function are displayed in Fig. 4. All models were adjusted for age, MVPA, and dietary protein intake, and muscle function models were additionally adjusted for subcutaneous fat. Regarding body composition in the entire cohort (Fig. A4), progesterone was positively associated with ALM and ALM index (*p*<0.040 and 0.043, respectively) and FEI, progesterone, and FAI were positively associated with body fat percentage (*p*<0.001, p=0.003, 0.005, respectively). FEI and FAI were positively associated with intramuscular fat (*p*<0.024 and 0.040, respectively). In contrast, the TT/E2 ratio was negatively associated with ALM index (*p*=0.025), and thigh muscle CSA (*p*=0.004).

**Figure 4.**
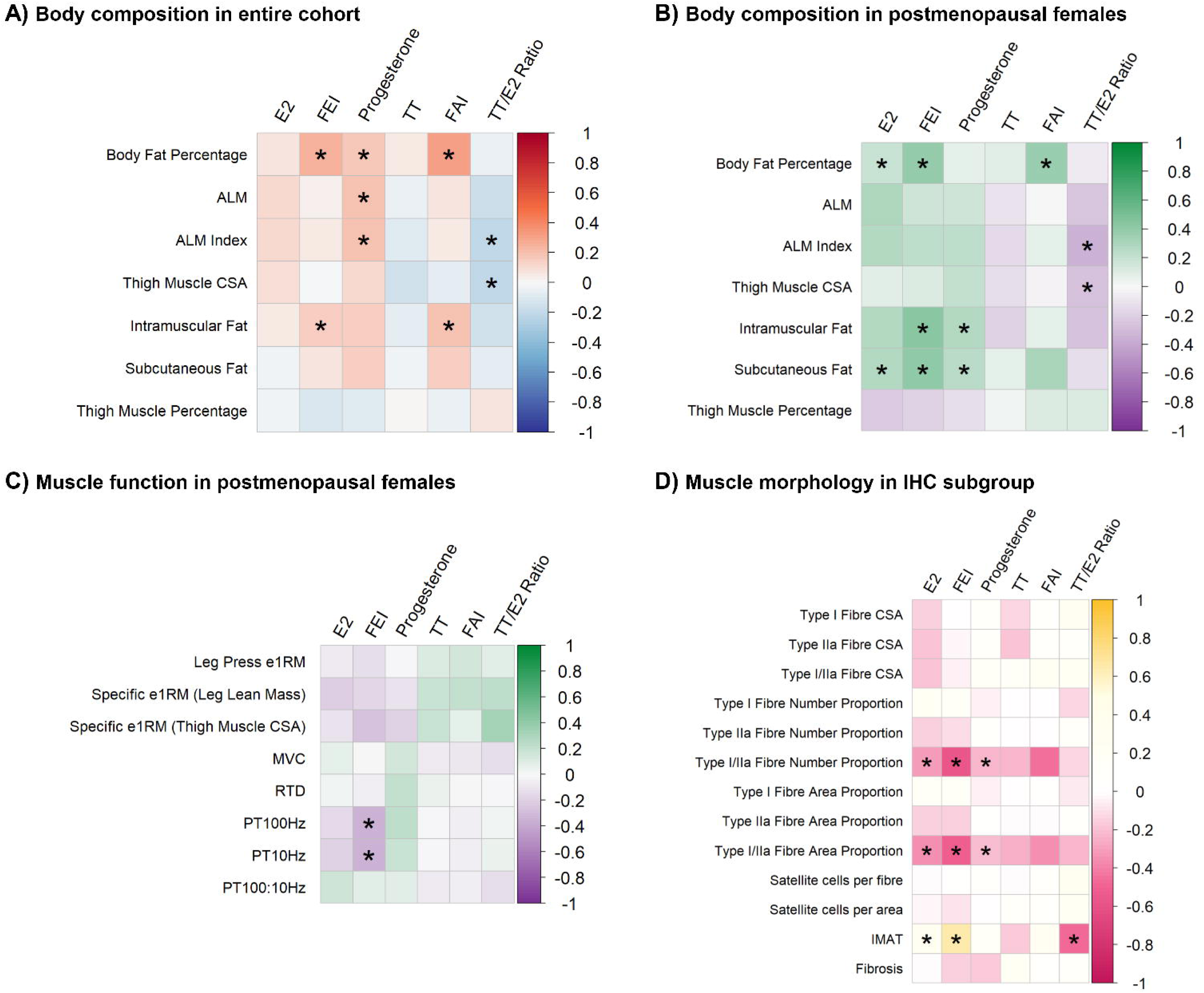
Linear associations between circulating sex hormones and muscle outcomes. A) Associations between sex hormones and body composition outcomes in entire cohort (n=81-96). B) Associations between sex hormones and body composition in postmenopausal females (n=37-45). C) Associations between sex hormones and muscle function outcomes in postmenopausal females (n=27-38). Muscle function variables were not tested in entire cohort because testing occurred on different day of the menstrual cycle to sex hormone quantification. D) Associations between sex hormones and muscle fibre characteristics in the subset of participants that underwent immunohistochemistry (IHC) analysis (n=30-36). Colour scale represents the standardised coefficient (β). Asterisk (*) denotes statistical significance (*p*<0.05). All models were adjusted for age, moderate-vigorous physical activity (MVPA), and protein intake. Muscle strength models were additionally adjusted for thigh subcutaneous fat. Specific e1RM was normalised to leg lean mass (kg/kg) and thigh muscle cross-sectional area (kg/cm^2^). All significant associations between circulating sex hormones and muscle outcomes have been plotted and are displayed in Supplementary Figures S1-4 on Figshare (https://doi.org/10.26187/deakin.30163165.v1). ALM: appendicular lean mass; CSA: cross-sectional area; e1RM: estimated one-repetition maximum; E2: oestradiol; FAI: free androgen index; FEI: free oestradiol index; IMAT: intermyofibrillar adipose tissue; MVC: maximal voluntary contraction; PT10Hz: peak potentiated twitch torque at 10 Hz; PT100Hz: peak potentiated twitch torque at 100 Hz; RTD: rate of torque development; TT: total testosterone.

In postmenopausal females only, E2, FEI, and FAI were positively associated with body fat percentage (*p*=0.021, 0.005, 0.006, respectively), FEI and progesterone were positively associated with thigh intramuscular (*p*=0.014 and 0.048, respectively) and subcutaneous fat (*p*=0.048 and 0.043, respectively). E2 was positively associated with subcutaneous fat (*p*=0.009) and the TT/E2 ratio was negatively associated with ALM index (*p*=0.030) and thigh muscle percentage (*p*=0.024; Fig. 4B). Associations between sex hormones and muscle function could only be investigated in postmenopausal females, where FEI was negatively associated with PT_10Hz_ (*p*=0.025) and PT_100Hz_ (*p*=0.013; Fig. 4C). Total testosterone was not associated with any body composition or muscle function measure (*p*>0.05).

Associations between circulating sex hormones and muscle morphology outcomes are displayed in Fig. 4D. All models were adjusted for age, MVPA, and dietary protein intake. E2, FEI and progesterone were all negatively associated with the proportion of hybrid muscle fibres by both number (*p*=0.009, 0.002, 0.005, respectively) and area (*p*=0.001, p<0.001, p=0.005, respectively). E2 and FEI were positively associated with IMAT percentage (*p*=0.048, p<0.001, respectively), while TT/E2 was negatively associated with it (*p*=0.026). There were no significant associations between sex hormone levels and type I or IIa fibre size or proportion, satellite cell number, or fibrosis (*p*>0.05).

All significant associations between circulating sex hormones and muscle outcomes have been plotted and are displayed in Supplementary Figures S1-4 on Figshare (https://doi.org/10.26187/deakin.30163165.v1).

### Female muscle transcriptome across the lifespan

In a sub-sample of 57 females stratified by decade of age, age was linearly associated on a log scale with the expression levels of 1603 genes in skeletal muscle (Fig. 5). The top-ranked pathways that were activated with age were related to the immune response and cellular adhesion, while the top-ranked pathways that were supressed with age were related to mitochondrial respiration.

**Figure 5.**
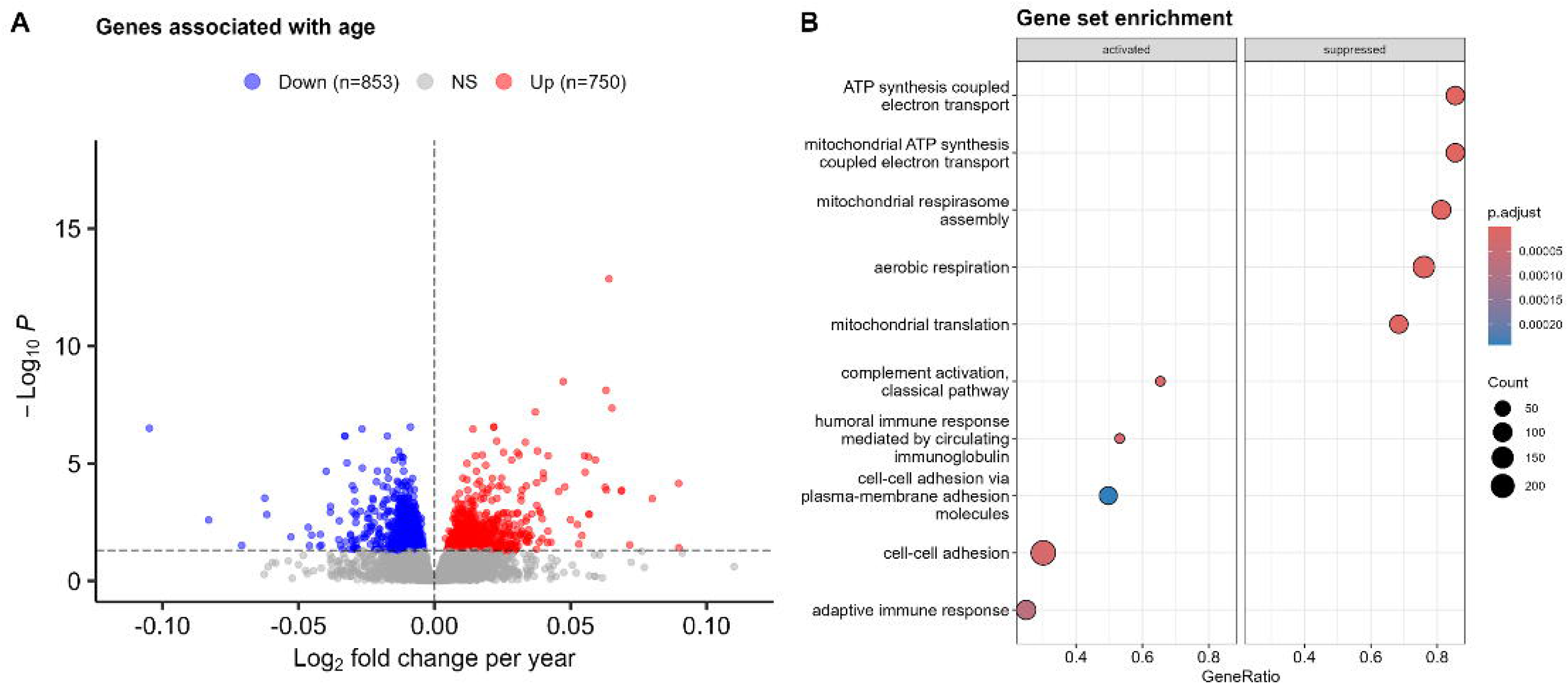
Linear associations between age and muscle gene expression (n=57). A) Volcano plot of DESeq2 investigating effect of age on gene expression (expression ∼ batch + age). Red and blue markers indicate genes that are up- and down-regulated with age, respectively. B) Gene set enrichment analysis of significant genes using Gene Ontology biological pathways, showing the most activated and most suppressed pathways with ageing. Adjusted *p*-value threshold < 0.05 using Benjamini-Hochberg correction. NS: non-significant.

The Likelihood Ratio Test identified 3255 genes that were significantly different across individual age groups (adjusted *p*<0.05), which were grouped into 37 visible clusters that exhibited unique patterns of expression (Fig. 6). Eight clusters of interest were selected based on the number of genes they contained, the pattern of expression, and the biological relevance of the pathway enrichment based on existing literature. Cluster 3 (227 genes; Fig. 7A) and 6 (367 genes; Fig. 7C) demonstrate an overall increase in gene expression across age groups and were related to RNA transcription and extracellular matrix organisation, respectively. Genes in cluster 4 (248 genes; Fig. 7B) and 16 (122 genes; Fig. 7G) show an overall decline in expression across the lifespan and relate to the ubiquitin proteosome system and cell cycle, respectively. Several clusters (13, 15, and 20; Fig. 7E-F & H) related to aerobic respiration and mitochondrial complex I biogenesis demonstrated an increase in expression between 18-29 and 30-39 years, followed by a decrease in expression across the later decades. Genes relating to autophagy and mitophagy were represented in cluster 7 (74 genes; Fig. 7D), also showing an inverted U-shape pattern of expression across the lifespan.

**Figure 6.**
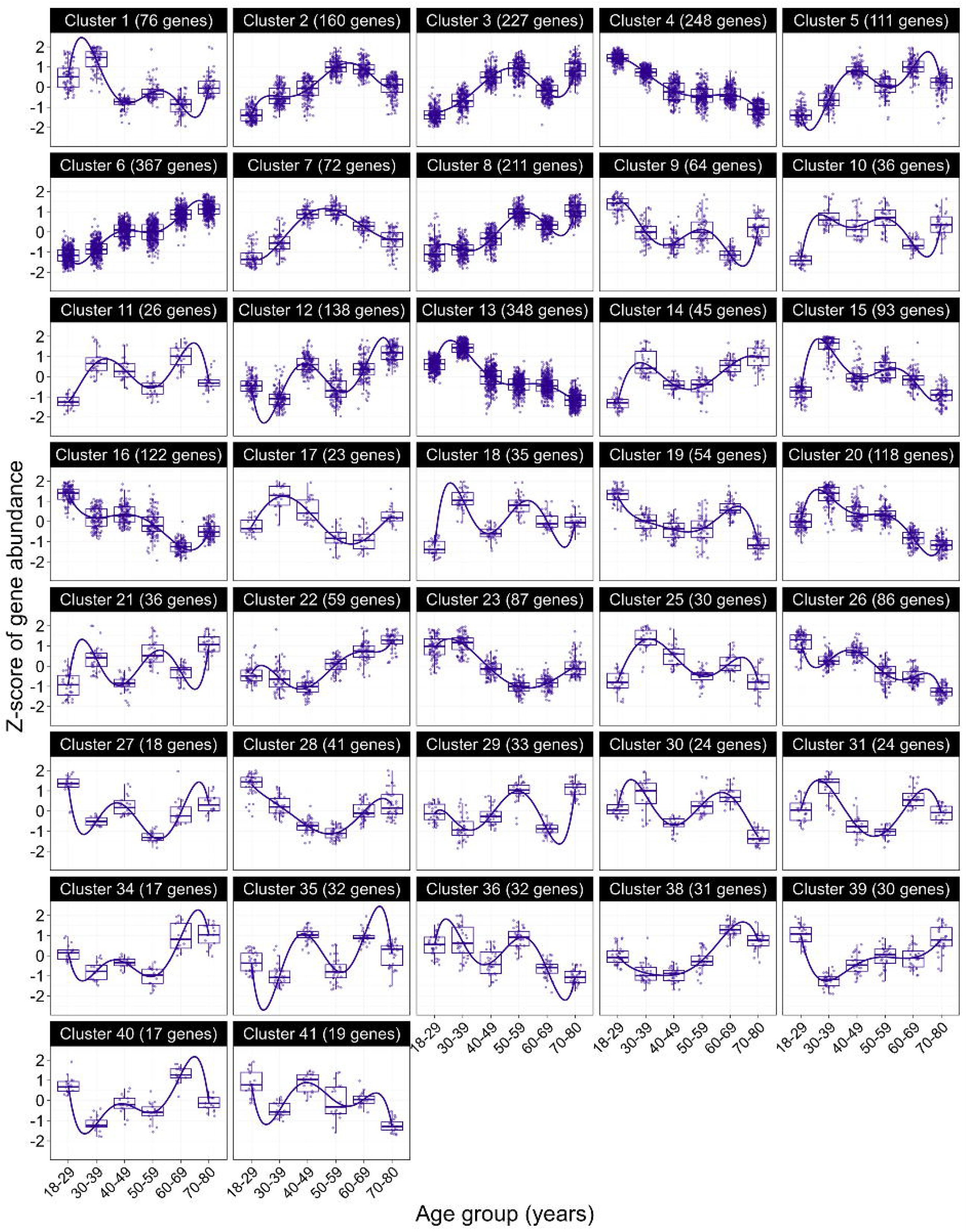
Muscle gene expression patterns across the lifespan (n=57). All gene clusters showing unique patterns of expression in genes differentially expressed across individual age groups.

**Figure 7.**
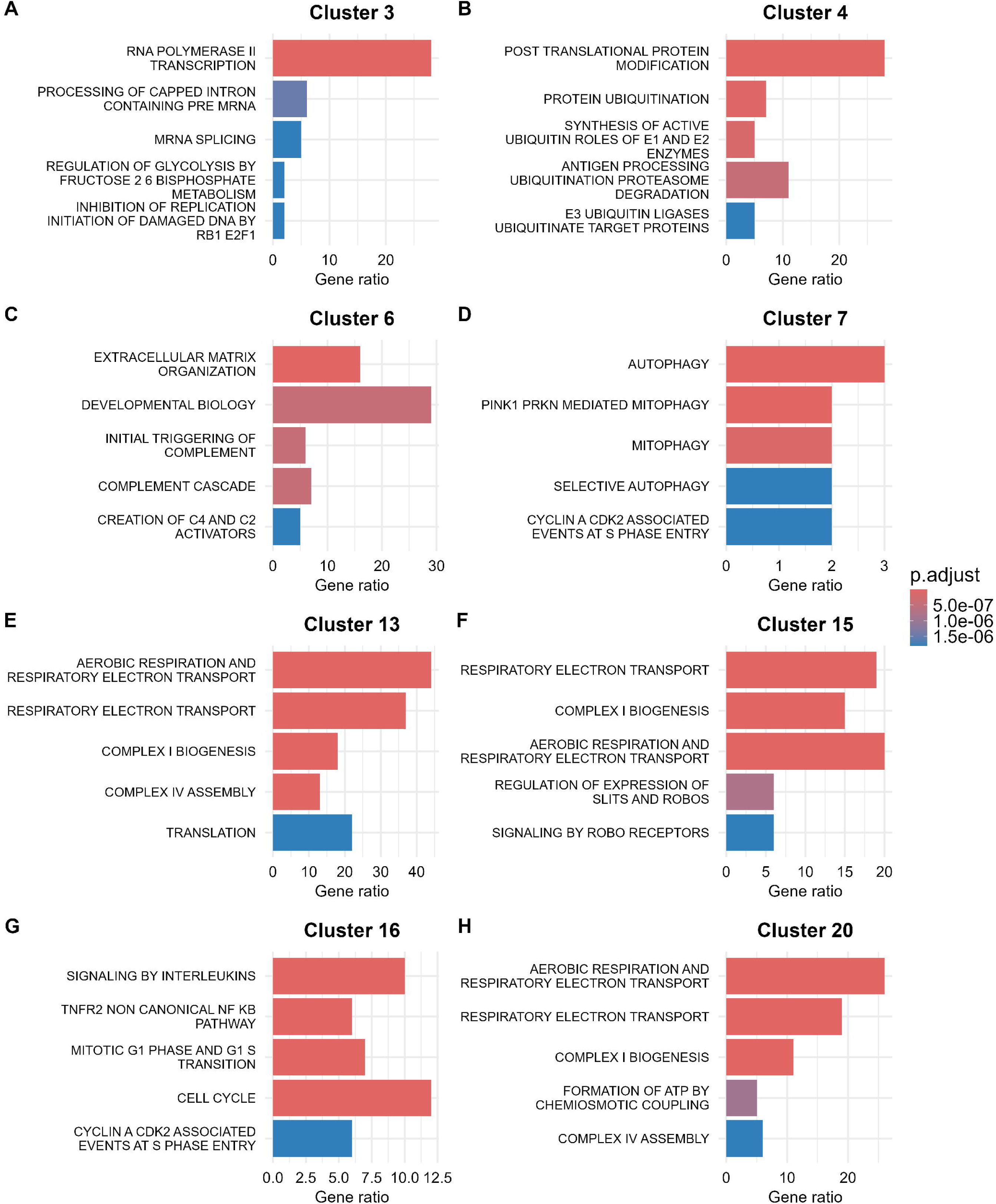
Overrepresentation analysis of muscle signalling pathways using the Reactome gene set in the top eight clusters of interest (n=57). Adjusted *p*-value threshold < 0.05 using Benjamini-Hochberg correction.

Associations between circulating sex hormones and gene expression are displayed in Fig. 8. Circulating E2 and FEI were associated with the expression of 14 (Fig, 8A) and 19 genes (Fig, 8B), respectively (adjusted p<0.05). Testosterone and FAI were associated with the expression of six genes each (Fig. 8C-D), and progesterone was associated with the expression of nine genes (Fig. 8E; adjusted p<0.05). A list of the genes significantly associated with each hormone is shown in Table A4.

**Figure 8.**
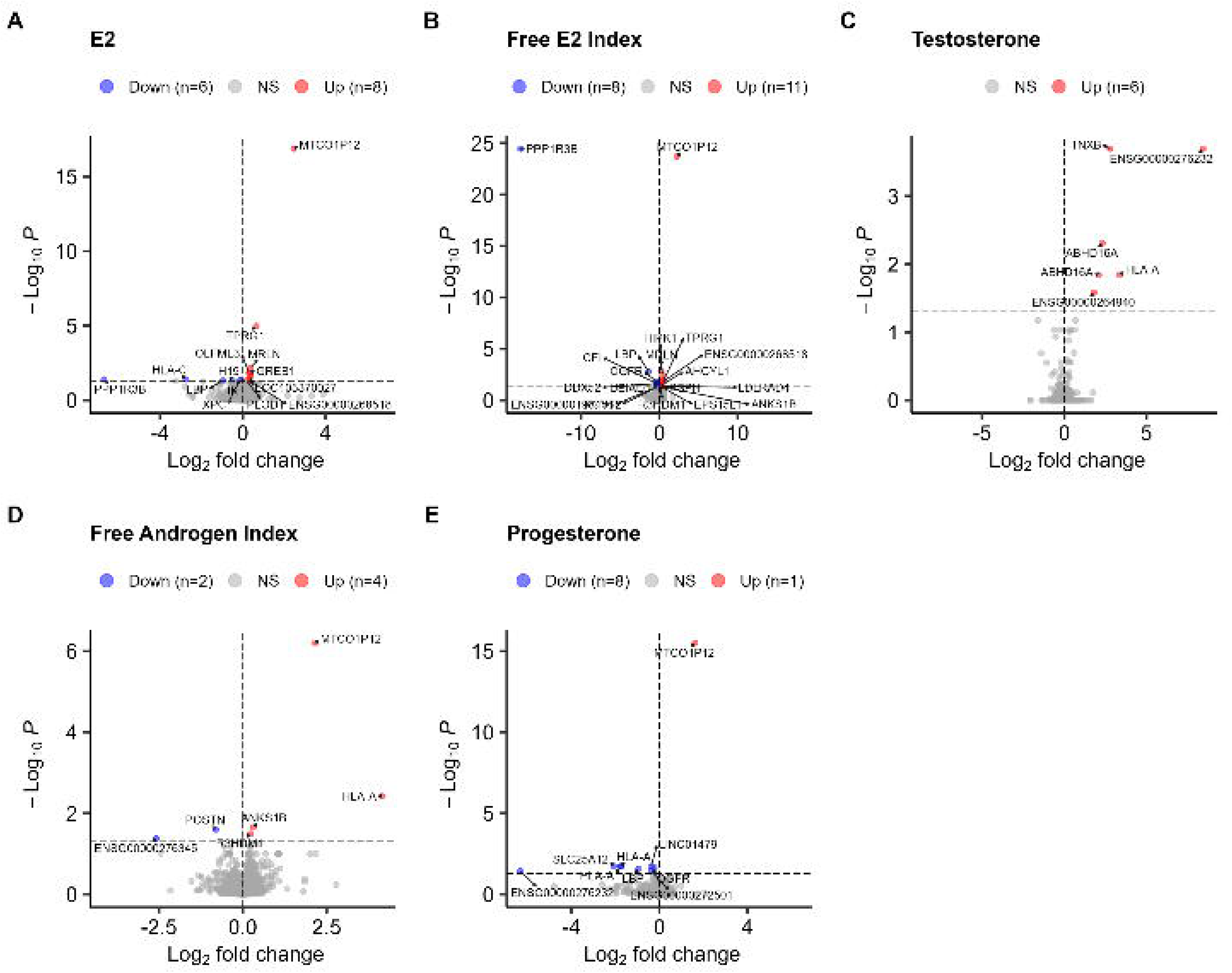
Linear associations between circulating sex hormones and muscle gene expression (n=57). A-E) Volcano plot of DESeq2 investigating effect of sex hormones on gene expression (expression ∼ batch + hormone concentration). Red and blue markers indicate genes that are up- and down-regulated with increasing sex hormone concentrations, respectively. Unidentified genes are labelled with the Ensemble gene ID. Adjusted *p*-value threshold < 0.05 using Benjamini-Hochberg correction. NS: non-significant.

Sixty-four TFs were enriched in the 511 genes that were upregulated in pre- versus post-menopausal females (*p<*0.01; Fig. 9A). In contrast, only three TFs were enriched in the 517 genes downregulated in pre- versus post-menopausal females (*p<*0.01; Fig. 9B). The oestrogen receptor (ESR1) was the 20^th^ most enriched TF in premenopausal females. The androgen (AR) and progesterone (PGR) receptors were not significantly enriched in any group.

**Figure 9.**
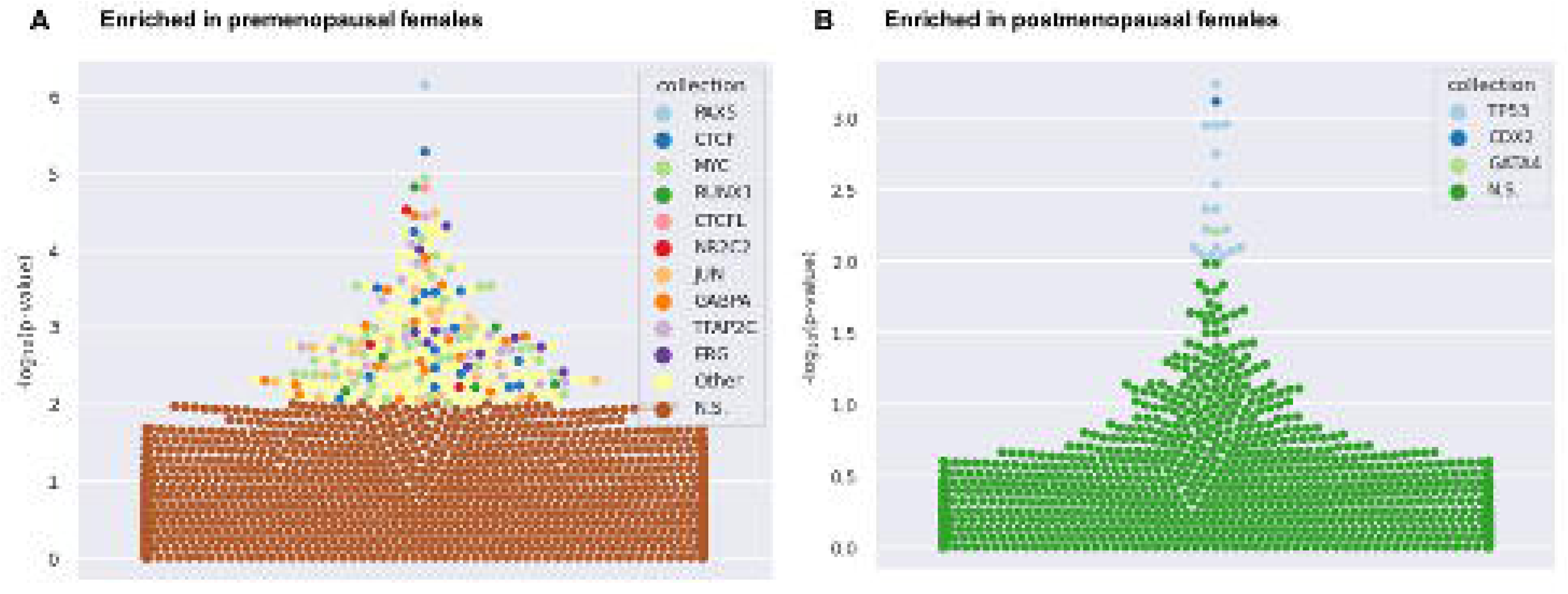
Transcription factor enrichment with ageing and menopause (n=57). A) Bee swarm plot showing enrichment of TFBS in genes that were significantly upregulated in pre- versus post-menopausal females. B) Bee swarm plot showing enrichment of TFBS in genes that were significantly upregulated in post- versus pre-menopausal females. The top 10 most enriched TFs are shown by different coloured markers, and the number of markers in each colour represent the number of datasets for each TF. The significance threshold was *p* < 0.01 using Fisher’s exact tests.

Finally, we deconvoluted our bulk muscle tissue transcriptome results using MuSiC deconvolution. Muscle tissue samples comprised a mix of RNA from muscle, stromal, endothelial, immune cells, or erythrocytes. There were no significant differences in the proportions of RNA originating from any of these cell types across age groups (*p*>0.05, Fig. 10A). Likewise, there was no difference in the proportion of RNA content originating from each muscle cell type (slow, fast, smooth, or satellite cell, *p*>0.05, Fig. 10B).

**Figure 10.**
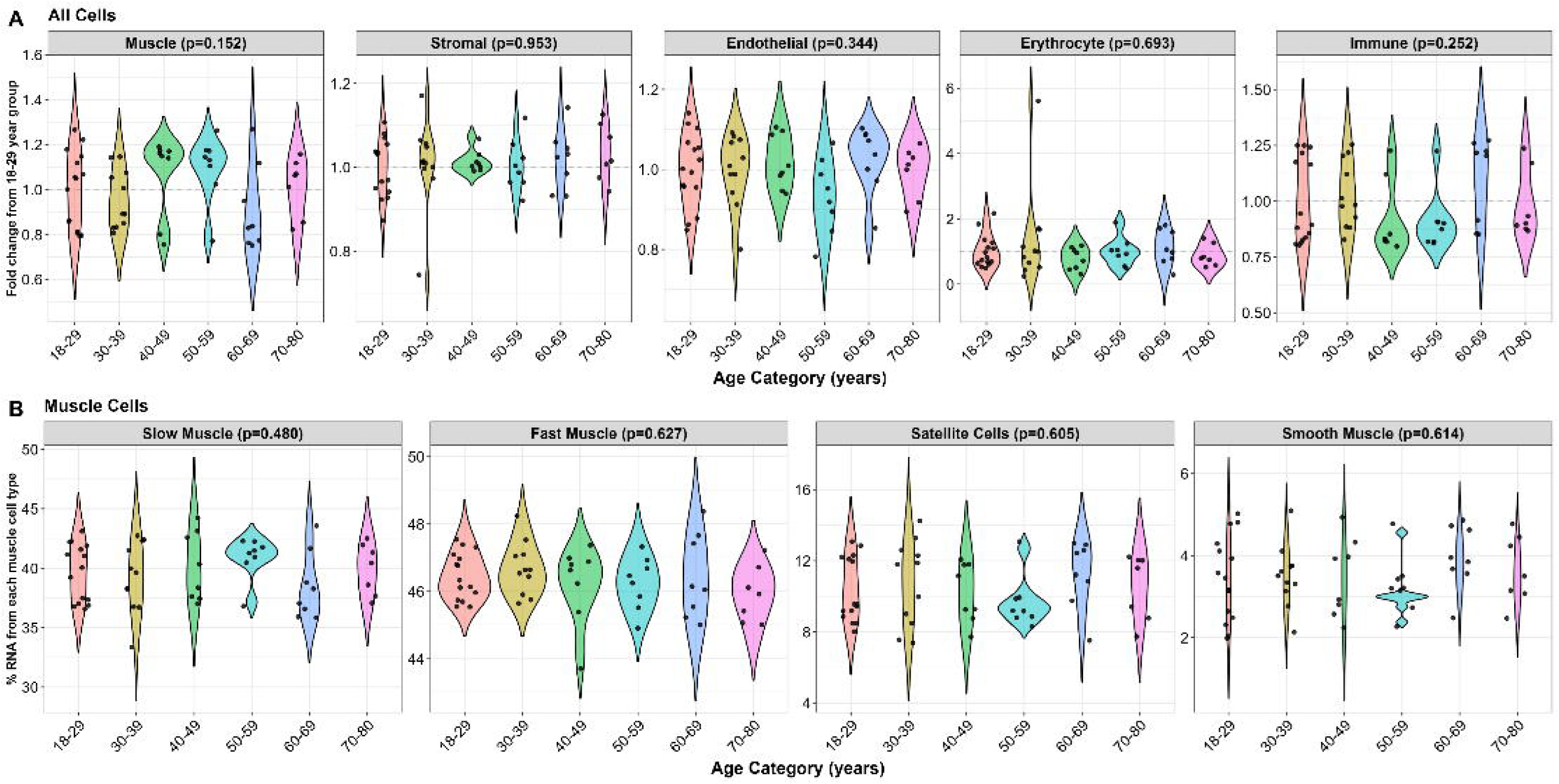
Sequenced muscle RNA originating from each cell type (n=57). A) Proportion of RNA originating from each cell type, normalised to the 18-29 year-old group. B) Percentage of each muscle cell type across individual age groups. Differences between age groups measured by one-way ANOVA with a statistical significance threshold of *p*<0.05.

## Discussion

This is the first study to map skeletal muscle ageing across the entire adult female lifespan at the whole-body, whole-muscle, cellular, and molecular level in tightly controlled conditions.

### Linear declines in muscle mass and strength across the female lifespan

This study demonstrates a linear reduction in lean mass (ALM and thigh muscle CSA), muscle quality (intramuscular fat and thigh muscle percentage), and muscle strength (absolute and specific leg press strength) in 96 females aged 18-80 years after adjusting for MVPA and protein intake. Across the lifespan, we report a loss of ALM and leg press e1RM by 0.4kg and 7.1kg per decade, respectively. Compared to the 18–29-year-olds, thigh muscle CSA was 27.9% lower in the 70–80-year-olds, and they were 38.6% weaker in the leg press. Other studies have demonstrated an acceleration in the loss of both muscle mass (Janssen *et al*., 2002) and strength (Haynes *et al*., 2020) around the age of menopause, but we did not observe a change in the rate of age-related muscle wasting at any time across the lifespan, as all models were best fit with a linear regression equation. This is contrast to the results from the analysis of the neuromuscular outcomes of the same cohort, reported in O’Bryan *et al*. (2025), where we found that voluntary and evoked quadriceps function (MVC, RTD, and PT) declines more rapidly beyond the onset of the menopausal transition (40-47 years) and that the decline in endogenous circulating sex hormones is a significant contributor post-menopause. While these outcomes reflect knee extensor function, the leg press is a compound exercise that involves multiple muscle groups, so the discrepancy could be explained by different rates of muscle strength loss with age or menopause across different compartments of the thigh.

### Circulating E2 and progesterone are associated with muscle mass and function across the lifespan

Previous analysis from ∼170,000 females in the UK BioBank highlighted that postmenopausal status was significantly associated with lower muscle mass compared to premenopausal status (Fieldsend *et al*., 2025), which aligns with several studies showing a significant loss of ALM and/or muscle CSA across the menopausal transition (Pöllänen *et al*., 2011, 2015; Nyberg *et al*., 2017; Park *et al*., 2020; Juppi *et al*., 2020). However, isolating the effects of declining circulating sex hormones from age is challenging, as they occur simultaneously. Here we first showed that, after adjusting for age and influential lifestyle factors (MVPA and protein intake), circulating E2, FEI and progesterone concentrations across the adult lifespan are significantly associated with absolute and relative ALM. These findings support previous work by our group (Critchlow *et al*., 2025), which utilised the Baltimore Longitudinal Study of Aging dataset (Shock, 1984) to demonstrate that after adjusting for age and other confounding factors, total circulating E2 and FEI were positively associated with muscle mass across the entire adult female lifespan. Together this suggests that the decline in endogenous ovarian hormone concentrations compounds the age-related loss of muscle mass across the female lifespan.

Across the entire cohort (18-80 years of age) we also found that greater FEI was strongly associated with infiltration of adipose tissue between the muscle fibres, which is a negative indicator of muscle quality. In addition, postmenopausal E2, FEI, and progesterone concentrations were positively associated with higher body fat and greater thigh subcutaneous and intramuscular fat. While this may seem contradictory, adipose tissue can produce sex hormones, therefore greater fat mass may lead to higher circulating and bioavailable hormone concentrations (Cui *et al*., 2013). These findings suggest that postmenopausal females could be particularly sensitive to changes in extra-gonadal production of sex hormones because their basal circulating levels are substantially lower than menstruating females. However, the direction and exact nature of this relationship cannot be confirmed from this analysis, therefore further investigation is required.

In addition, this study found a negative association between FEI and peak potentiated twitch torque of the quadriceps. The direction of these associations is unexpected as previous evidence suggests that changes in circulating E2 from high to low concentrations negatively impact muscle function. For example, a reduction in FEI across 4-6 years of ageing was longitudinally associated with a loss of handgrip strength in 83 females over 50 years old (Critchlow *et al*., 2025). However, a recent systematic review showed a consistent lack of cross-sectional association between serum or urinary E2 and muscle strength in postmenopausal females (Critchlow *et al*., 2023), likely because in this population, E2 concentrations are low and thereafter remain stable. While we adjusted our muscle function models to account for differences in subcutaneous fat, the present study’s results may be explained by the increase in intramuscular adipose tissue with increasing E2 concentrations, which may worsen overall muscle quality and attenuate the twitch signal (Tomazin *et al*., 2011).

Finally, while 18.3% of peri- and post-menopausal participants were currently using a form of HRT, this was not associated with differences in measures of muscle mass or function, suggesting HRT does not minimise age-related muscle wasting in this cohort. This result should however be interpreted with caution as the effects of HRT were not a primary outcome of the study, potentially leading to a lack of statistical power to detect the true effects of HRT. The literature regarding HRT’s effects in postmenopausal female muscle is conflicting (Critchlow *et al*., 2023), with some studies showing positive effects on muscle strength (Greising *et al*., 2009) but not mass (Javed *et al*., 2019). In addition, the effectiveness of exogenous oestrogen and progesterone supplementation likely depends on the composition, dose and duration of treatment (Critchlow *et al*., 2023), as well as the time since menopause (Park *et al*., 2020) and the accurate reporting of these variables.

### Circulating testosterone is not associated with muscle mass or strength across the lifespan

In contrast to E2 and progesterone, we observed no significant associations between total circulating testosterone and any measure of body composition, muscle morphology, or muscle strength. This adds to the recent, growing body of evidence showing no association between total circulating testosterone and muscle mass or strength in pre- (Alexander *et al*., 2021, 2024) or post-menopausal (Gower & Nyman, 2000; Sipilä *et al*., 2006; Rariy *et al*., 2011; Bann *et al*., 2015) females. FAI has previously been positively and more strongly linked with muscle mass and strength than total testosterone in pre- (Alexander *et al*., 2021, 2024) and post-menopausal (Kong *et al*., 2019) females, likely because it more accurately represents the bioavailable fraction of testosterone that is able to enter the muscle cell, but we did not observe any relationship between FAI and muscle mass or function. Together these findings indicate that circulating fractions of testosterone across the lifespan are less relevant in the regulation of female skeletal muscle mass and function across ageing compared to E2 and progesterone.

It is also important to consider the interactions between the different sex hormones as they share a common biosynthesis pathway (Cui *et al*., 2013). For example, the TT/E2 ratio represents the conversion of testosterone to E2 by the enzyme aromatase (Swislocki & Eisenberg, 2024). Here we report significant negative associations between the TT/E2 ratio and lean mass and muscle size in both the entire cohort (18-80 years of age) and in postmenopausal females only. Additionally, we previously reported that a greater TT/E2 ratio was associated with smaller thigh muscle CSA and worse hamstring strength in females across the lifespan (24-89 years of age) (Critchlow *et al*., 2025). Together these findings indicate that females with greater conversion of testosterone to E2 (higher aromatase activity) exhibit better muscle mass irrespective of age. In contrast to E2 and progesterone, circulating testosterone concentrations remain relatively stable across and beyond menopause (Handelsman *et al*., 2016). It is therefore possible that higher extra-gonadal conversion of testosterone to E2 in the later decades of life can act as a compensatory mechanism to counteract the cessation of ovarian E2 production, limiting age-related muscle wasting, although further investigation is required to confirm this.

### The proportion of hybrid muscle fibres in the *vastus lateralis* increases with age

This study demonstrated age-related atrophy of type I fibres by size across the female lifespan, but there was no association between ageing and the proportion of pure type I or type IIa fibres in the *vastus lateralis*, neither by number nor area. In addition, after deconvolution of our bulk RNA-Seq results, we observed no significant change in the amount of RNA originating from slow or fast muscle cells with age relative to the 18–29-year-old group average. Originating from early studies in mostly male subjects, there is a widespread perception that ageing leads to the preferential atrophy of type II fibres, leading to an increased proportion of type I fibres within the muscle (Larsson *et al*., 1978). However, other studies have often shown conflicting results, either reporting increased (Horwath *et al*., 2025), decreased (Klitgaard *et al*., 1990), or no change (Lim *et al*., 2019) to the type I fibre proportion in old versus young individuals. These differences likely arise from confounding factors such as sex, physical activity levels, or location of biopsy, which can all significantly alter the expression of myosin heavy chain (MyHC) isoforms (Wilson *et al*., 2012; Haizlip *et al*., 2015; Van De Casteele *et al*., 2024). A recent meta-analysis of 27 studies by Lee *et al*. (2024) found an overall age-related increase in MyHC I protein expression and a reduction in MyHC II fibre CSA, but when stratified by sex, the females (21% of participants) showed no significant change to fibre type proportions with age. Females have a greater proportion of type I fibres at baseline compared to males and demonstrate unique fibre-type responses to other muscle wasting conditions such as cancer cachexia (Anoveros-Barrera *et al*., 2019), heart failure (Wood *et al*., 2024), and disuse (Callahan *et al*., 2015). This strongly suggests that mechanisms of age-related and pathological muscle wasting are sex-specific, further emphasising the importance of female-only analyses. However, while this study reports an age-related reduction in the size of type I, but not IIa, fibres, previous studies have sometimes demonstrated the opposite pattern (Grosicki et al., 2022) or no changes (Sato *et al*., 1984; Essén-Gustavsson & Borges, 1986; Coggan *et al*., 1992; Miller *et al*., 2013) when comparing young and old females. Reasons for these discrepancies are unknown, but our study is the first to map muscle fibre size and type across every decade of the adult female lifespan while controlling for important lifestyle factors (MVPA and protein intake) and may therefore overcome some of the limitations of earlier studies.

Despite no age-related change in the proportion of pure type I and IIa fibres, we observed a significant age-related increase in the proportion of hybrid fibres co-expressing MyHC I and IIa isoforms. Hybrid fibres possess the contractile properties intermediate to the MyHC isoforms they express, and have often been shown to increase with age (Klitgaard *et al*., 1990; D’Antona, 2003; St-Jean-Pelletier *et al*., 2017), with the proportion of type I/IIa fibres ranging from 6-8% in young (20-30 years) and 15-30% in older males (65-90 years). Here, we observed much smaller values, with an average of 2% in 18–29-year-olds, and 3% in 70-80-year-olds, suggesting they do not likely play a large physiological role within ageing female muscle. Interestingly, circulating E2 (total and free) and progesterone were negatively associated with the proportion of hybrid I/IIa fibres in the *vastus lateralis*, but not pure type I or IIa fibres, after adjusting for age, MVPA, and protein intake. This suggests that ovarian hormones may regulate the co-expression of MyHC isoforms, but not individual fibre types. Potential targets of E2 and/or progesterone and their receptors may include factors that mediate fibre-type-specific transcription, such as calcineurin or nuclear factor of activated T cells (NFAT) (Chin *et al*., 1998).

While previous studies have demonstrated a loss of satellite cells in old males and females compared to their young counterparts (Kadi *et al*., 2004; Verdijk *et al*., 2007), we found no effect of age on satellite cell number in the *vastus lateralis* of females aged 18 to 80 years after controlling for physical activity and protein intake. We also found no association between circulating sex hormones and satellite cell number, which contrasts with findings from Collins *et al*. (2019) who found a positive association in five females across the menopausal transition. Likewise, several studies report that ovariectomised rodents have fewer satellite cells than intact counterparts (Frechette *et al*., 2015; Collins *et al*., 2019), and that their function is significantly impaired (Kitajima & Ono, 2016). Further research would therefore be beneficial in humans to determine whether E2 may moderate the response to muscle damage after exercise or injury.

### The muscle transcriptome exhibits unique patterns of expression across the female lifespan

This study is the first to extensively map muscle gene expression across the entire adult female lifespan. Using DEseq2, we found 1603 genes that were significantly and linearly regulated with age (as a continuous variable) on the log scale in the *vastus lateralis* of healthy females, 750 and 853 of which were up- and down-regulated across the lifespan, respectively. Several studies have recently compared the *vastus lateralis* muscle transcriptome in young (18-30 years) and old (>65 years) individuals (De Jong *et al*., 2023; Pataky *et al*., 2023; Huang *et al*., 2024), but these dichotomised groups omit the ability to detect changes in gene expression across the mid-decades of life, which is particularly crucial in females who undergo dramatic changes to their sex hormone profile within this period. In addition, using likelihood ratio testing we found 3255 genes whose expression changes (linearly or non-linearly) across individual decades of age. Within these genes, cluster analysis found 37 unique patterns of expression, highlighting the need to consider non-linear patterns of muscle gene expression across the lifespan.

Gene set enrichment analysis of age-associated genes and overrepresentation analysis of the age-group gene clusters consistently highlighted that pathways relating to mitochondrial function and aerobic respiration were suppressed with age. Specifically, downregulated genes were enriched in pathways regulating mitochondrial ATP synthesis via the electron transport chain and mitochondrial biogenesis and translation. Huang *et al*. (2024) also reported female enrichment of age-induced DEGs in pathways related to the electron transport chain and cellular metabolism. Faulty ATP production and mitochondrial protein translation can contribute to cellular stress via an increased release of reactive oxygen species (ROS) and an accumulation of cell debris, leading to oxidative stress and apoptosis (Mansouri *et al*., 2006; Chabi *et al*., 2008). In combination, clusters four and seven show that processes responsible for degrading damaged proteins, such as the ubiquitin proteosome system, autophagy, and mitophagy are supressed in the later decades, potentially causing further damage to the cell and leading to a loss of muscle mass and quality (Wohlgemuth *et al*., 2010).

Our findings also show that the top activated pathways with age relate to immune signalling, cellular adhesion, and extracellular matrix organisation. Deconvolution of our RNA sequencing results confirmed the presence of resident immune cells, although the proportion of RNA originating from these cells did not change with age. Nevertheless, increased inflammatory signalling with age (‘inflammaging’) is well understood to negatively contribute to age-related muscle wasting via tumour necrosis factor alpha (TNF-α), which activates pathways of protein degradation (Sishi & Engelbrecht, 2011). Pro-inflammatory interleukin-6 can also act via signal transducer and activator of transcription 3 (STAT3) to suppress myogenic lineage progression and exhaust the satellite cell pool (Sala & Sacco, 2016). Cellular adhesion is also a crucial feature of cellular signalling that allows communication and structural support between cells and the extracellular matrix (Gumbiner, 1996). The extracellular environment relies on highly coordinated signalling patterns between multiple cell types, including immune cells, fibroblasts, and progenitor cells, to be able to respond appropriately to stress or injury (Chinvattanachot *et al*., 2024). Dysregulation of this process with ageing may contribute to the disruption of proliferation and differentiation signals, leading to satellite cell senescence and an accumulation of fibrosis and fat within the muscle (Chinvattanachot *et al*., 2024). Together, these findings highlight relevant potential targets for therapeutic interventions to combat the deterioration of female muscle signalling with age.

### Circulating sex hormones are weakly associated with the female muscle transcriptome

We next aimed to determine the associations between circulating sex hormone concentrations and the resting muscle transcriptome across the lifespan. We found only a small number of genes (n = 6 to 19) that were significantly associated with E2, progesterone, and testosterone concentrations after adjusting for false discovery rate (adjusted *p*-value < 0.05). In contrast, Pataky *et al*. (2023) found 1,131 gene transcripts that were significantly associated with circulating E2 in young (18-30 years) and old (65-80 years) females after adjusting for age. This stark difference may be explained by their use of multivariate regression analysis, while the present study used generalised linear models to account for non-normal distribution and overdispersion of the data, both of which are characteristic features of RNA-sequencing counts data (Conesa *et al*., 2016).

Here we found that total, but not free, E2 was positively associated with GREB1 expression, a cofactor for ER transcriptional activity which is upregulated upon ER binding to oestrogen response elements (EREs) on target genes (Hodgkinson *et al*., 2018). While GREB1 has been studied in the context of breast and ovarian cancer (Hodgkinson *et al*., 2018), its specific role within skeletal muscle remains unknown, but these findings suggest it may be a regulator of muscle E2 signalling. Interestingly, expression of ESR1 (the gene that encodes the ER) was not significantly associated with circulating E2 or age, but there was significant enrichment of ESR1 binding sites in premenopausal females versus postmenopausal females, suggesting greater transcription of ER target genes when circulating E2 is higher.

We also report that E2 and FEI were positively associated with MRLN expression, the gene that encodes myoregulin. This protein moderates calcium reuptake into the sarcoplasmic reticulum via inhibition of sarcoplasmic/endoplasmic reticulum calcium ATPase 1 (SERCA1) (Anderson *et al*., 2015). SERCA pumps regulate muscle contraction and relaxation by transporting Ca^2+^ back into the sarcoplasmic reticulum (SR) from the cytosol (Xu & Van Remmen, 2021). Genetic knock-out of myoregulin significantly improves running performance in young male mice, but further investigation is required to understand its potential role in female muscle (Anderson *et al*., 2015). Also of note is MTCO1P12, a pseudogene whose expression is positively and most strongly associated with E2 and progesterone concentrations. While it does not code a functional protein, it could play a regulatory role via genomic interactions, although further research is needed to understand its relationship with female sex hormones.

### Study limitations

While this study has presented novel patterns of muscle ageing across the female lifespan, it does not come without limitations. Firstly, while every effort was made to complete all testing in the early-follicular phase of the menstrual cycle where relevant, this was not achieved in 20-24% of participants due to the unpredictability of cycles, masking of bleeding by hormonal contraception (e.g. IUD), and availability of testing dates. Variability of menstrual cycle phase at visit two therefore explains some of the variation in mean circulating E2 and progesterone across the younger decades of age (Table 2). While we found no inter-individual effect of menstrual cycle phase on our muscle function outcomes (Table 3), future studies should track menstrual cycles at least two months before participation commences using a cycle calendar (Elliott-Sale *et al*., 2021).

Secondly, the lack of non-linear declines in muscle mass or strength with age was unexpected and contrasts with other studies demonstrating an acceleration in age-related muscle wasting during the later decades (Janssen *et al*., 2002; Haynes *et al*., 2020) including our own (O’Bryan *et al*., 2025).

Robust and unbiased linearity testing consistently found a better fit of linear models, but this may be explained by an insufficient number of participants around the age of menopause. While each decade of age was similarly represented in the cohort, only four participants were classified as perimenopausal (as diagnosed by a medical practitioner), although determination of menopausal stage can be difficult at this age due to masking of menstrual cycle patterns by hormonal contraceptives. Previous analysis of the same cohort found non-linear declines in neuromuscular function with age, suggesting that the study was sufficiently powered to detect non-linear changes, but future studies should aim to maximise recruitment around the age of menopause to ensure sufficient sensitivity to detect any potential acceleration of muscle mass and strength loss.

Similarly, while we were sufficiently powered to detect changes in our primary outcomes (muscle mass and strength) across the lifespan, this study includes several subgroup analyses, including transcriptomics (n=57) and immunohistochemistry (n=36), where statistical power is reduced from the overall sample. Sensitivity analysis of our transcriptomic data determined our study to be sufficiently powered to reliably detect moderate effect sizes at FDR < 0.05. However, as this is the first study to present histological analysis of female muscle across the female lifespan, our immunohistochemistry results should be viewed as exploratory and provide a foundation for future, more well-powered, studies. Finally, we elected to include female participants who were using a range of hormonal contraceptives and hormone replacement therapies to ensure generalisability of findings to wider populations. This meant that we were not statistically powered to investigate the individual effects of each form of exogenous hormone supplementation on our outcome measures, although this was not a primary aim of the study.

## Conclusion

This study is the first to map the trajectory of female muscle ageing at the whole-body, whole-muscle, and cellular level, while tightly controlling for important lifestyle factors such as MVPA, protein intake, and menstrual cycle. Female ageing is associated with a loss of muscle mass and leg press strength, a reduction in muscle fibre size, and an increase in hybrid muscle fibres and fibrosis. We identified unique patterns of muscle gene expression across the adult female lifespan, with upregulation of immune signalling pathways and downregulation of pathways relating to mitochondrial function. Independent of age, circulating E2 and progesterone, but not testosterone, concentrations are positively associated with lean mass and negatively associated with hybrid muscle fibres. Together these findings suggest that female-specific factors of ageing, such as ovarian hormone fluctuations, should be considered when developing therapeutic interventions to combat age-related muscle wasting. However, longitudinal and interventional studies are required to confirm the causal nature of the relationship between sex hormones and female muscle ageing.

## Data Availability

All data produced in the present study are available upon reasonal request to the authors.

https://github.com/acritchlow/FAMe_Analysis

## Additional Information

### Data availability statement

The R code for analysis can be found at https://github.com/acritchlow/FAMe_Analysis. Raw data are available upon request. RNA sequencing counts have been deposited on the Gene Expression Omnibus (GEO) platform under accession number GSE303107.

### Author contributions

A.J.C. was involved in acquisition, analysis and interpretation of data, and drafting and revising the manuscript for intellectual content. D.H. was involved in conception and experimental design, acquisition, analysis and interpretation of data and drafting and revising the manuscript for intellectual content. S.O.B. was involved in conception and experimental design, acquisition and analysis of data, and revising the manuscript for intellectual content. M.S. was involved in analysis of data and revising the manuscript for intellectual content. R.M.W. was involved in acquisition of data and revising the manuscript for intellectual content. V.E. was involved in acquisition of data and revising the manuscript for intellectual content. K.V.B. was involved in acquisition of data and revising the manuscript for intellectual content. R.P.W. was involved in acquisition of data and revising the manuscript for intellectual content. A.G. was involved in acquisition of data and revising the manuscript for intellectual content. C.S.F. was involved in acquisition of data and revising the manuscript for intellectual content. D.S. was involved in interpretation of data and revising the manuscript for intellectual content. S.L. was involved in conception and experimental design, interpretation of data and drafting and revising the manuscript for intellectual content and funded the study.

## Acknowledgments

The authors would like to thank the participants who volunteered their time and Ms Briana Gatto for her role in managing participant recruitment and coordination during the early phases of the study.

## Appendix

**Table A1.** Characteristics of sample subset that underwent muscle fibre histological analysis.

| Variable | Overall<br>N = 36 | 18-29<br>N = 6 | 30-39<br>N = 6 | 40-49<br>N = 6 | 50-59<br>N = 6 | 60-69<br>N = 6 | 70-80<br>N = 6 |
| --- | --- | --- | --- | --- | --- | --- | --- |
| **Body composition** |  |  |  |  |  |  |  |
| Weight (kg) | 68.9 (12.3) | 65.5 (11.4) | 63.7 (13.1) | 70.9 (11.3) | 71.8 (17.4) | 70.8 (9.1) | 70.3 (13.5) |
| Height (m) | 1.65 (0.06) | 1.64 (0.07) | 1.64 (0.06) | 1.67 (0.07) | 1.70 (0.05) | 1.65 (0.05) | 1.60 (0.03) |
| BMI (kg.m<sup>-2</sup>) | 25.3 (4.7) | 24.2 (4.9) | 23.8 (5.5) | 25.5 (3.3) | 24.8 (5.8) | 26 (4.8) | 27.3 (4.8) |
| Body fat (%) | 33.6 (10.1) | 29.1 (8.1) | 29.8 (12) | 33.1 (9.8) | 32.3 (10.0) | 39.1 (10.1) | 38.4 (10.5) |
| Total body fat mass (kg) | 23.0 (10.7) | 18.6 (8.3) | 19.4 (12) | 22.5 (9.6) | 23.4 (14.2) | 27.1 (10.1) | 27 (11.5) |
| Total body lean mass (kg) | 42.9 (4.3) | 43.6 (6.3) | 41.7 (3.0) | 45.5 (2.9) | 45.2 (4.4) | 40.6 (3.6) | 40.4 (2.1) |
| ALM (kg) | 19.2 (2.3) | 20.4 (2.9) | 18.7 (1.7) | 20.2 (1.4) | 19.9 (2.7) | 18.1 (1.8) | 17.5 (1.7) |
| ALM index (kg.m<sup>-2</sup>) | 7.0 (0.7) | 7.5 (1.0) | 6.9 (0.5) | 7.3 (0.4) | 6.9 (0.9) | 6.7 (0.7) | 6.8 (0.6) |
| Bone mineral content (kg) | 2.4 (0.4) | 2.3 (0.4) | 2.3 (0.2) | 2.6 (0.2) | 2.6 (0.4) | 2.2 (0.4) | 2.2 (0.4) |
| **Muscle size and composition** |  |  |  |  |  |  |  |
| Thigh muscle CSA (cm<sup>2</sup>) | 95.4 (17.1) | 110 (13.1) | 105.8 (13.2) | 104.3 (12.2) | 87 (17.8) | 86.9 (12.9) | 78.9 (10.1) |
| Intramuscular fat (cm<sup>2</sup>) | 3.9 (2.6) | 3.1 (2.1) | 2.2 (1.7) | 3.4 (1.9) | 4.1 (1.4) | 5.6 (4.2) | 5.0 (2.7) |
| Subcutaneous fat (cm<sup>2</sup>) | 71.5 (29.6) | 69.5 (38.7) | 64.9 (33.2) | 72.6 (25.6) | 57.8 (7.1) | 85.4 (39.2) | 75.7 (25.9) |
| Relative thigh muscle (%) | 56.2 (24.2) | 57.2 (10.7) | 58.1 (10.4) | 54.0 (6.9) | 79.8 (57.7) | 46.3 (9.9) | 45.9 (6.6) |
| Femur bone density (g.cm<sup>-3</sup>) | 775.3 (59.7) | 799.6 (49) | 812 (16.3) | 813.3 (45.5) | 755.3 (44) | 776.2 (70.7) | 697.9 (30) |
| Muscle fibre type proportion by number (%) |  |  |  |  |  |  |  |
| *Type I* | 52.5 (15.6) | 53.6 (11.1) | 46.1 (20.2) | 54.3 (22.8) | 49.0 (9.1) | 56.8 (15.7) | 55.2 (15.1) |
| *Type IIa* | 45.9 (15.7) | 44.5 (10.1) | 53.0 (20.5) | 45.2 (23.0) | 48.5 (9.4) | 41.8 (15.9) | 42.5 (15.2) |
| *Type I/IIa hybrid* | 2.4 (2.3) | 2.2 (2.2) | 1.3 (1.5) | 0.9 (0.9) | 4.6 (3.5) | 2.4 (1.7) | 3.3 (1.8) |
| Muscle fibre type proportion by area (%) |  |  |  |  |  |  |  |
| *Type I* | 49.7 (16.2) | 55.7 (13.1) | 43.7 (19.5) | 51.9 (22.2) | 43.3 (9.5) | 54.1 (17.1) | 49.2 (16.1) |
| *Type IIa* | 47.0 (16.4) | 40.5 (11.4) | 53.3 (19.7) | 45.2 (23.0) | 52.9 (9.6) | 42.5 (17.6) | 47.3 (16.3) |
| *Type I/IIa hybrid* | 2.3 (2.2) | 1.7 (1.3) | 1.3 (1.7) | 0.8 (0.9) | 4.4 (2.7) | 2.8 (2.4) | 3.1 (2.0) |
| Mean muscle fibre CSA (µm<sup>2</sup>) |  |  |  |  |  |  |  |
| *Type I* | 4,148.3 (1,116.2) | 5,565.9 (991.1) | 4,259.7 (1,064.3) | 4,169.2 (733.6) | 4,047.3 (1,103.6) | 3,520.3 (543.8) | 3,327.5 (913.1) |
| *Type IIa* | 4,674.4 (1,072.2) | 4,569.8 (896.7) | 4,807.9 (989.7) | 4,534.2 (811.6) | 5,596.3 (1,529.1) | 3,986.8 (628.2) | 4,551.3 (1,099.1) |
| *Type I/IIa hybrid* | 3,751.0 (1,039.1) | 4,595.6 (1,710.0) | 3,388.6 (697.7) | 3,606.0 (596.5) | 4,039.2 (889.1) | 3,866.1 (725.2) | 2,900.8 (805.8) |
| *All fibres* | 4,248.0 (943.5) | 4,952.0 (889.2) | 4,394.2 (1,046.9) | 4,244.9 (697.3) | 4,530.6 (1,149.3) | 3,651.4 (540.1) | 3,715.2 (872.9) |
| IMAT (%) | 1.8 (2.1) | 0.6 (0.2) | 1.4 (1.1) | 2.1 (2.0) | 2.0 (1.5) | 3.8 (4.5) | 1.2 (0.6) |
| Fibrosis (%) | 9.3 (3.0) | 7.7 (2.1) | 9.4 (2.8) | 6.5 (1.9) | 9.4 (2.2) | 10.2 (4.1) | 11.9 (2.1) |
| Satellite cell number |  |  |  |  |  |  |  |
| *Per fibre* | 0.13 (0.05) | 0.14 (0.06) | 0.16 (0.03) | 0.12 (0.05) | 0.16 (0.06) | 0.10 (0.02) | 0.12 (0.04) |
| *Per unit of area* | 15.5 (5.5) | 13.62 (6.6) | 15.98 (4.3) | 14.13 (4.5) | 17.37 (5.1) | 14.11 (7.3) | 17.82 (5.2) |
| **Muscle strength** |  |  |  |  |  |  |  |
| Leg press e1RM (kg) | 91.4 (27.3) | 118.0 (35.1) | 97.0 (27.4) | 103.0 (14.7) | 80.9 (20.4) | 74.8 (13.5) | 69.2 (7.4) |
| Specific leg press e1RM |  |  |  |  |  |  |  |
| *Normalised to leg lean mass*<br>(kg.kg<sup>-1</sup>) | 6.2 (1.7) | 7.8 (2.4) | 6.7 (1.7) | 6.7 (0.6) | 5.3 (1.4) | 5.4 (0.9) | 5.3 (0.9) |
| *Normalised to thigh muscle CSA*<br>(kg.cm<sup>-2</sup>) | 0.93 (0.19) | 1.08 (0.29) | 0.90 (0.19) | 1.00 (0.21) | 0.87 (0.17) | 0.86 (0.11) | 0.88 (0.1) |
| **Circulating sex hormones** |  |  |  |  |  |  |  |
| Total E2 (pmol/L) | 196.8 (195) | 206.6 (130.9) | 268 (215.1) | 455 (253.5) | 114.9 (117.1) | 84.9 (20.3) | 61.8 (14.3) |
| Free E2 index (FEI) | 0.44 (0.54) | 0.44 (0.39) | 0.64 (0.6) | 1.07 (0.83) | 0.22 (0.14) | 0.18 (0.07) | 0.1 (0.07) |
| Total progesterone (nmol/L) | 11.6 (19.9) | 9.9 (14.1) | 14.3 (21.1) | 36.4 (34) | 3.9 (2.8) | 3.5 (3.1) | 2.5 (0.6) |
| Total testosterone (nmol/L) | 0.82 (0.35) | 0.95 (0.41) | 0.88 (0.25) | 0.71 (0.2) | 0.77 (0.41) | 0.70 (0.4) | 0.92 (0.41) |
| Free androgen index (FAI) | 1.7 (1.10) | 2.2 (1.70) | 1.8 (1.0) | 1.5 (0.8) | 1.8 (1.1) | 1.5 (1.0) | 1.6 (1.2) |
| TT/E2 ratio | 7.7 (6.2) | 5.9 (3.4) | 4.5 (2.8) | 2.3 (1.9) | 9.3 (6.1) | 8.4 (4.4) | 15.7 (7.7) |
| **Reproductive health characteristics** |  |  |  |  |  |  |  |
| Menopausal status |  |  |  |  |  |  |  |
| *Premenopausal* | 18 / 36 (50%) | 7 / 7 (100%) | 5 / 5 (100%) | 5 / 6 (83%) | 1 / 6 (17%) | - | - |
| *Perimenopausal* | 2 / 36 (6%) | - | - | 1 / 6 (17%) | 1 / 6 (17%) | - | - |
| *Postmenopausal* | 16 / 36 (44%) | - | - | - | 4 / 6 (67%) | 6 / 6 (100%) | 6 / 6 (100%) |
| Hormonal contraception |  |  |  |  |  |  |  |
| *None* | 10 / 36 (28%) | 2 / 7 (29%) | 2 / 5 (40%) | 4 / 6 (67%) | 2 / 6 (33%) | - | - |
| *OCP* | 2 / 36 (6%) | 1 / 7 (14%) | 1 / 5 (20%) | - | - | - | - |
| *IUD* | 8 / 36 (22%) | 4 / 7 (57%) | 2 / 5 (40%) | 2 / 6 (33%) | - | - | - |
| *Implant* | 0 / 36 (0%) | - | - | - | - | - | - |
| *N/A (postmenopausal)* | 16 / 36 (44%) | - | - | - | 4 / 6 (67%) | 6 / 6 (100%) | 6 / 6 (100%) |
| Hormonal replacement therapy |  |  |  |  |  |  |  |
| *Currently using* | 4 / 36 (11%) | - | - | 1 / 6 (17%) | - | 2 / 6 (33%) | 1 / 6 (17%) |
| *Not currently using* | 14 / 36 (39%) | - | - | - | 5 / 6 (83%) | 4 / 6 (67%) | 5 / 6 (83%) |
| *N/A (premenopausal)* | 18 / 36 (50%) | 7 / 7 (100%) | 5 / 5 (100%) | 5 / 6 (83%) | 1 / 6 (17%) | - | - |
| Menstrual cycle phase at muscle biopsy |  |  |  |  |  |  |  |
| *Early follicular* | 8 / 36 (22%) | 3 / 6 (50%) | 2 / 6 (33.3%) | 2 / 6 (33.3%) | 1 / 6 (16.7%) | - | - |
| *Late follicular* | 2 / 36 (6%) | - | 2 / 6 (33.3%) | - | - | - | - |
| *Ovulation* | 1 / 36 (3%) | - | - | 1 / 6 (16.7%) | - | - | - |
| *Mid-luteal* | 5 / 36 (14%) | 1 / 6 (16.7%) | 1 / 6 (16.7%) | 3 / 6 (50%) | - | - | - |
| *Unknown* | 2 / 36 (6%) | 1 / 6 (16.7%) | - | - | 1 / 6 (16.7%) | - | - |
| *N/A* | 18 / 36 (50%) | 1 / 6 (16.7%) | 1 / 6 (16.7%) | - | 4 / 6 (66.7%) | 6 / 6 (100%) | 6 / 6 (100%) |
| **Lifestyle factors** |  |  |  |  |  |  |  |
| Dietary protein intake (g.kg<sup>-1</sup>.day<sup>-1</sup>) | 0.93 (0.19) | 1.08 (0.29) | 0.90 (0.19) | 1.00 (0.21) | 0.87 (0.17) | 0.86 (0.11) | 0.88 (0.10) |
| MVPA (mins.day<sup>-1</sup>) | 41.8 (34.7) | 35.5 (14.0) | 48.8 (21.0) | 54.8 (60.5) | 38.0 (16.5) | 48.0 (47.4) | 26.0 (36.3) |

Presented as mean (SD). ALM: appendicular lean mass; CSA: cross-sectional area; e1RM: estimated one-repetition maximum; E2: oestradiol; IMAT: intermyofibrillar adipose tissue; IUD: intrauterine device; MVPA: moderate to vigorous physical activity; OCP: oral contraceptive pill; TT: total testosterone.

**Table A2.** Characteristics of sample subset that underwent RNAseq analysis.

| Variable | Overall<br>N = 57 | 18-29<br>N = 15 | 30-39<br>N = 11 | 40-49<br>N = 8 | 50-59<br>N = 8 | 60-69<br>N = 8 | 70-80<br>N = 7 |
| --- | --- | --- | --- | --- | --- | --- | --- |
| Body composition |  |  |  |  |  |  |  |
| Weight (kg) | 68.7 (12.9) | 66.7 (15.0) | 69 (13.1) | 71.6 (7.6) | 66.9 (16.7) | 73.8 (10.0) | 65.3 (12.7) |
| Height (m) | 1.64 (0.07) | 1.65 (0.08) | 1.64 (0.07) | 1.65 (0.06) | 1.66 (0.07) | 1.65 (0.07) | 1.61 (0.04) |
| BMI (kg.m<sup>-2</sup>) | 25.4 (4.7) | 24.6 (5.2) | 25.7 (4.1) | 26.3 (3.7) | 24.3 (5.1) | 27.2 (5.9) | 25.2 (4.2) |
| Body fat (%) | 34.5 (9.0) | 31.8 (7.7) | 35.5 (9.4) | 34.4 (7.8) | 30.8 (9.4) | 39.7 (10.1) | 36.8 (9.7) |
| Total body fat mass (kg) | 23.3 (9.9) | 20.6 (8.9) | 24.3 (10.3) | 23.5 (8) | 20.9 (12.8) | 28.8 (10.7) | 24.0 (9.1) |
| Total body lean mass (kg) | 42.3 (5.7) | 42.6 (8.4) | 41.7 (4.5) | 45.1 (2.8) | 43.2 (5.0) | 42.2 (4.2) | 38.5 (4.1) |
| ALM (kg) | 18.7 (3) | 19.3 (4.5) | 18.5 (2.5) | 20.1 (1) | 19.0 (2.7) | 18.4 (2.0) | 16.3 (2.1) |
| ALM index (kg.m<sup>-2</sup>) | 6.9 (0.9) | 7.1 (1.4) | 6.9 (0.8) | 7.3 (0.3) | 6.9 (0.7) | 6.8 (0.6) | 6.3 (0.6) |
| Bone mineral content (kg) | 2.4 (0.4) | 2.4 (0.5) | 2.4 (0.3) | 2.5 (0.3) | 2.5 (0.4) | 2.1 (0.3) | 2.2 (0.3) |
| Muscle size and composition |  |  |  |  |  |  |  |
| Thigh muscle CSA (cm<sup>2</sup>) | 94.2 (19.4) | 101.6 (22.8) | 99.9 (16.9) | 105.9 (11.7) | 86.6 (10.6) | 90 (11.5) | 69.2 (15.0) |
| Intramuscular fat (cm<sup>2</sup>) | 4.1 (2.7) | 3.8 (3.3) | 2.9 (1.6) | 3.8 (2.6) | 3.9 (1.5) | 6.0 (3.5) | 5.2 (1.6) |
| Subcutaneous fat (cm<sup>2</sup>) | 74.0 (32.5) | 77.1 (38.0) | 73.8 (27.3) | 83.4 (35.2) | 56.1 (8.9) | 87.5 (43.6) | 60.5 (21.4) |
| Relative thigh muscle (%) | 51.3 (8.6) | 52.7 (8.8) | 53.1 (8.3) | 52.1 (10.4) | 53.9 (4.4) | 46.9 (11.6) | 46.9 (4.3) |
| Femur bone density (g.cm<sup>-3</sup>) | 777.2 (74.8) | 802.2 (63.2) | 796.8 (66.9) | 818.8 (42.3) | 800.6 (38.8) | 720 (99.1) | 690.6 (27.9) |
| Muscle strength |  |  |  |  |  |  |  |
| Leg press e1RM (kg) | 87.1 (26.5) | 100.8 (33.1) | 93.3 (25) | 95.6 (18.3) | 79.9 (15) | 74.0 (13.7) | 59.4 (14.5) |
| Specific leg press e1RM |  |  |  |  |  |  |  |
| *Normalised to leg lean mass (kg.kg<sup>-1</sup>)* | 6.0 (1.3) | 6.7 (1.5) | 6.5 (1.3) | 6.2 (1) | 5.6 (1.1) | 5.4 (0.8) | 4.7 (0.8) |
| *Normalised to thigh muscle CSA (kg.cm<sup>-2</sup>)* | 0.9 (0.1) | 1.0 (0.2) | 0.9 (0.1) | 1.0 (0.1) | 0.9 (0.1) | 0.8 (0.1) | 0.8 (0.1) |
| Circulating sex hormones |  |  |  |  |  |  |  |
| Total E2 (pmol/L) | 214.4 (221.9) | 185.0 (166.1) | 384.0 (299.6) | 394.2 (241.4) | 132.4 (117.2) | 75.1 (10.1) | 58.5 (7.3) |
| Free E2 index (FEI) | 0.58 (0.96) | 0.70 (1.45) | 1.18 (1.07) | 0.76 (0.69) | 0.26 (0.16) | 0.18 (0.09) | 0.12 (0.11) |
| Total progesterone (nmol/L) | 11.1 (19.2) | 10.5 (19) | 17.3 (22.5) | 26.9 (30.5) | 4.2 (2.7) | 2.8 (0.6) | 2.3 (0.9) |
| Total testosterone (nmol/L) | 0.82 (0.39) | 0.94 (0.36) | 0.96 (0.47) | 0.78 (0.21) | 0.76 (0.41) | 0.63 (0.37) | 0.68 (0.48) |
| Free androgen index (FAI) | 2.0 (1.6) | 2.5 (2.2) | 2.9 (1.8) | 1.5 (0.9) | 1.7 (1.0) | 1.4 (1.0) | 1.3 (0.9) |
| TT/E2 ratio | 7.0 (5.6) | 7.2 (3.7) | 3.7 (2.8) | 3.7 (3.4) | 8.4 (6.7) | 8.2 (3.8) | 12.4 (9.4) |
| Reproductive health characteristics |  |  |  |  |  |  |  |
| Menopausal status |  |  |  |  |  |  |  |
| *Premenopausal* | 32 / 57 (56%) | 15 / 15 (100%) | 11 / 11 (100%) | 5 / 8 (63%) | 1 / 8 (13%) | - | - |
| *Perimenopausal* | 3 / 57 (5%) | - | - | 2 / 8 (25%) | 1 / 8 (13%) | - | - |
| *Postmenopausal* | 22 / 57 (39%) | - | - | 1 / 8 (13%) | 6 / 8 (75%) | 8 / 8 (100%) | 7 / 7 (100%) |
| Hormonal contraception |  |  |  |  |  |  |  |
| *None* | 24 / 57 (42%) | 9 / 15 (60%) | 7 / 11 (64%) | 6 / 8 (75%) | 2 / 8 (25%) | - | - |
| *OCP* | 4 / 57 (7%) | 3 / 15 (20%) | 1 / 11 (9%) | - | - | - | - |
| *IUD* | 6 / 57 (11%) | 2 / 15 (13%) | 3 / 11 (27%) | 1 / 8 (13%) | - | - | - |
| *Implant* | 1 / 57 (2%) | 1 / 15 (7%) | - | - | - | - | - |
| *N/A (postmenopausal)* | 22 / 57 (39%) | - | - | 1 / 8 (13%) | 6 / 8 (75%) | 8 / 8 (100%) | 7 / 7 (100%) |
| Hormonal replacement therapy |  |  |  |  |  |  |  |
| *Currently using* | 6 / 57 (11%) | - | - | 2 / 8 (25%) | 1 / 8 (13%) | 1 / 8 (13%) | 2 / 7 (29%) |
| *Not currently using* | 19 / 57 (33%) | - | - | 1 / 8 (13%) | 6 / 8 (75%) | 7 / 8 (88%) | 5 / 7 (71%) |
| *N/A (premenopausal)* | 32 / 57 (56%) | 15 / 15 (100%) | 11 / 11 (100%) | 5 / 8 (63%) | 1 / 8 (13%) | - | - |
| Menstrual cycle phase at muscle biopsy |  |  |  |  |  |  |  |
| *Early follicular* | 13 / 57 (23%) | 9 / 15 (60%) | 2 / 11 (18.2%) | 1 / 8 (12.5%) | 1 / 8 (12.5%) | - | - |
| *Late follicular* | 2 / 57 (4%) | - | 2 / 11 (18.2%) | - | - | - | - |
| *Ovulation* | 4 / 57 (7%) | - | 3 / 11 (27.3%) | 1 / 8 (12.5%) | - | - | - |
| *Mid-luteal* | 7 / 57 (12%) | 2 / 15 (13.3%) | 2 / 11 (18.2%) | 3 / 8 (37.5%) | - | - | - |
| *Unknown* | 4 / 57 (7%) | - | 1 / 11 (9.1%) | 2 / 8 (25%) | 1 / 8 (12.5%) | - | - |
| *N/A* | 27 / 57 (47%) | 4 / 15 (26.7%) | 1 / 11 (9.1%) | 1 / 8 (12.5%) | 6 / 8 (75%) | 8 / 8 (100%) | 7 / 7 (100%) |
| **Lifestyle factors** |  |  |  |  |  |  |  |
| Dietary protein intake (g.kg<sup>-1</sup>.day<sup>-1</sup>) | 1.3 (0.45) | 1.38 (0.6) | 1.56 (0.47) | 1.31 (0.37) | 1.37 (0.33) | 1.09 (0.15) | 1.1 (0.5) |
| MVPA (mins.day<sup>-1</sup>) | 38.8 (25.2) | 45.7 (25.1) | 44.4 (23.1) | 29 (14.6) | 46.3 (13.1) | 35.5 (33.6) | 20.3 (34.6) |

Presented as mean (SD). ALM: appendicular lean mass; CSA: cross-sectional area; e1RM: estimated one-repetition maximum; E2: oestradiol; IUD: intrauterine device; MVPA: moderate to vigorous physical activity; OCP: oral contraceptive pill; TT: total testosterone.

**Table A3.**
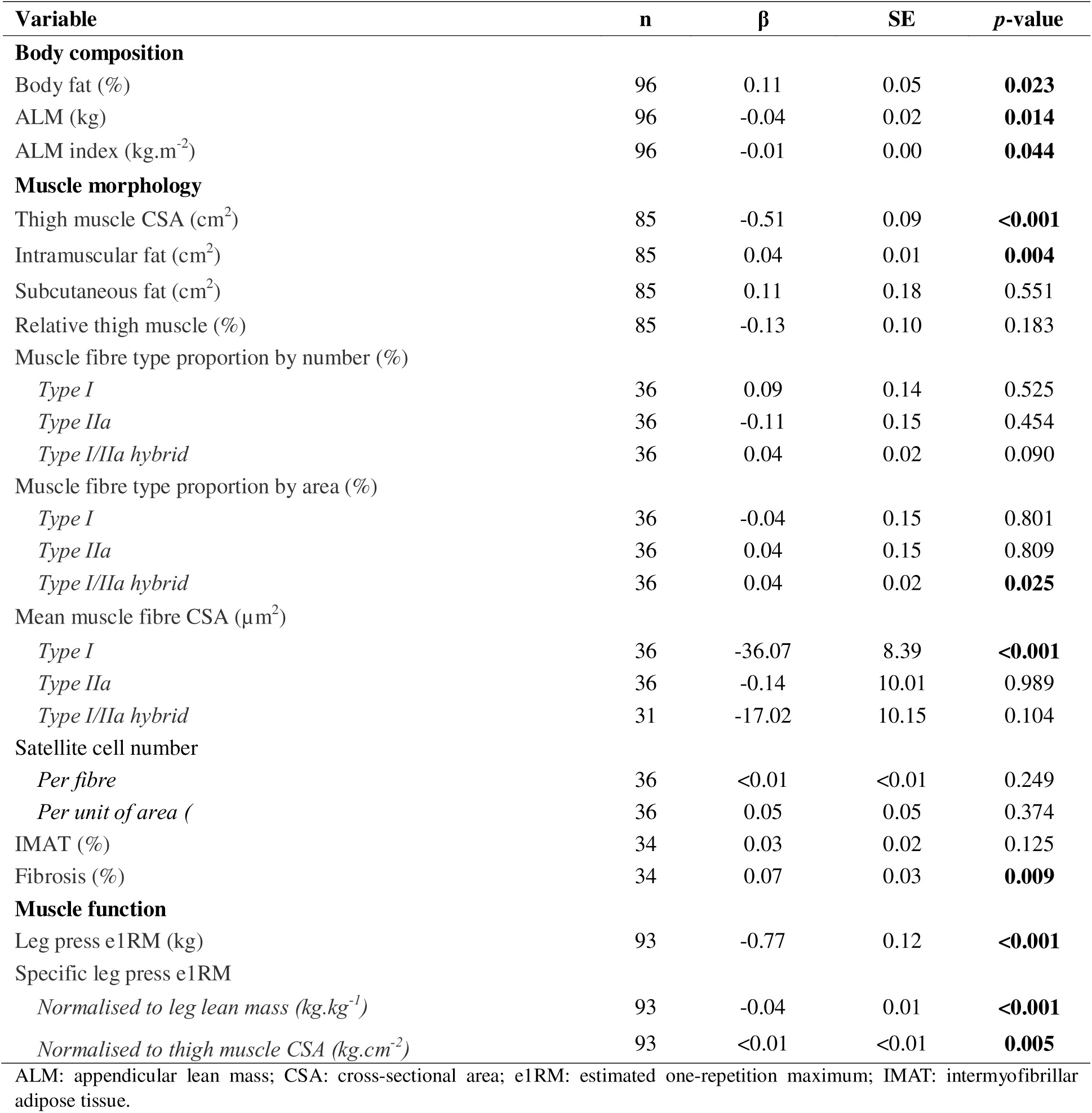
Unadjusted associations between age and female body composition, muscle morphology, and function.

| Variable | n | $\beta$ | SE | p-value |
| --- | --- | --- | --- | --- |
| **Body composition** |  |  |  |  |
| Body fat (%) | 96 | 0.11 | 0.05 | **0.023** |
| ALM (kg) | 96 | -0.04 | 0.02 | **0.014** |
| ALM index (kg.m<sup>-2</sup>) | 96 | -0.01 | 0.00 | **0.044** |
| **Muscle morphology** |  |  |  |  |
| Thigh muscle CSA (cm<sup>2</sup>) | 85 | -0.51 | 0.09 | **<0.001** |
| Intramuscular fat (cm<sup>2</sup>) | 85 | 0.04 | 0.01 | **0.004** |
| Subcutaneous fat (cm<sup>2</sup>) | 85 | 0.11 | 0.18 | 0.551 |
| Relative thigh muscle (%) | 85 | -0.13 | 0.10 | 0.183 |
| Muscle fibre type proportion by number (%) |  |  |  |  |
| Type I | 36 | 0.09 | 0.14 | 0.525 |
| Type IIa | 36 | -0.11 | 0.15 | 0.454 |
| Type I/IIa hybrid | 36 | 0.04 | 0.02 | 0.090 |
| Muscle fibre type proportion by area (%) |  |  |  |  |
| Type I | 36 | -0.04 | 0.15 | 0.801 |
| Type IIa | 36 | 0.04 | 0.15 | 0.809 |
| Type I/IIa hybrid | 36 | 0.04 | 0.02 | **0.025** |
| Mean muscle fibre CSA (μm<sup>2</sup>) |  |  |  |  |
| Type I | 36 | -36.07 | 8.39 | **<0.001** |
| Type IIa | 36 | -0.14 | 10.01 | 0.989 |
| Type I/IIa hybrid | 31 | -17.02 | 10.15 | 0.104 |
| Satellite cell number |  |  |  |  |
| Per fibre | 36 | <0.01 | <0.01 | 0.249 |
| Per unit of area ( | 36 | 0.05 | 0.05 | 0.374 |
| IMAT (%) | 34 | 0.03 | 0.02 | 0.125 |
| Fibrosis (%) | 34 | 0.07 | 0.03 | **0.009** |
| **Muscle function** |  |  |  |  |
| Leg press e1RM (kg) | 93 | -0.77 | 0.12 | **<0.001** |
| Specific leg press e1RM |  |  |  |  |
| Normalised to leg lean mass (kg.kg<sup>-1</sup>) | 93 | -0.04 | 0.01 | **<0.001** |
| Normalised to thigh muscle CSA (kg.cm<sup>-2</sup>) | 93 | <0.01 | <0.01 | **0.005** |

ALM: appendicular lean mass; CSA: cross-sectional area; e1RM: estimated one-repetition maximum; IMAT: intermyofibrillar adipose tissue.

**Table A4.** Genes significantly associated with circulating sex hormone concentration.

| Sex hormone | Genes |
| --- | --- |
| E2 | PLOD1, IK, OLFML3, LBP, XPC, TPRG1, GREB1, HLA-C, MRLN, MTCO1P12, LOC105370027, PPP1R3B, H19, ENSG00000268518 |
| FEI | R3HDM1, OGFR, UBIAD1, EPS15L1, LBP, PSPH, HIPK1, LDLRAD4, AHCYL1, ANKS1B, PCYT2, TPRG1, CFI, MRLN, MTCO1P12, DDX52, PPP1R3B, ENSG00000198406, ENSG00000268518 |
| Progesterone | OGFR, SLC25A12, LBP, HLA-A, MTCO1P12, LINC01479, ENSG00000272501, ENSG00000276232 |
| TT | ABHD16A, HLA-A, ABHD16A, TNXB, ENSG00000264940, ENSG00000276232 |
| FAI | R3HDM1, POSTN, ANKS1B, HLA-A, MTCO1P12, ENSG00000276345 |

Unidentified genes are labelled with their Ensemble ID. Adjusted p-value threshold < 0.05 using Benjamini-Hochberg correction. E2: oestradiol; FAI: free androgen index; FEI; free E2 index; TT: total testosterone.

## References

Alexander SE, Abbott G, Aisbett B, Wadley GD, Hnatiuk JA & Lamon S (2021). Total testosterone is not associated with lean mass or handgrip strength in pre-menopausal females. Sci Rep 11, 10226.

Alexander SE, Gatto B, Knowles OE, Williams RM, Fiebig KN, Jansons P, Della Gatta PA, Garnham A, Eynon N, Wadley GD, Aisbett B, Hiam D & Lamon S (2024). Bioavailable testosterone and androgen receptor activation, but not total testosterone, are associated with muscle mass and strength in females. J Physiol; DOI: 10.1113/JP286803.

Anderson DM, Anderson KM, Chang C-L, Makarewich CA, Nelson BR, McAnally JR, Kasaragod P, Shelton JM, Liou J, Bassel-Duby R & Olson EN (2015). A Micropeptide Encoded by a Putative Long Noncoding RNA Regulates Muscle Performance. Cell 160, 595–606.

Anoveros-Barrera A, Bhullar AS, Stretch C, Esfandiari N, Dunichand-Hoedl AR, Martins KJB, Bigam D, Khadaroo RG, McMullen T, Bathe OF, Damaraju S, Skipworth RJ, Putman CT, Baracos VE & Mazurak VC (2019). Clinical and biological characterization of skeletal muscle tissue biopsies of surgical cancer patients. J Cachexia Sarcopenia Muscle 10, 1356– 1377.

Bann D, Wu FCW, Keevil B, Lashen H, Adams J, Hardy R, Muniz G, Kuh D, Ben-Shlomo Y & Ong KK (2015). Changes in testosterone related to body composition in late midlife: Findings from the 1946 British birth cohort study. Obes Silver Spring Md 23, 1486–1492.

Barbosa M, Shiguemoto G, Tomaz L, Ferreira F, Rodrigues M, Domingues M, Souza Master M, Canevazzi G, Silva-Magosso N, Selistre-de-Araujo H & Perez S (2016). Resistance Training and Ovariectomy: Antagonic Effects in Mitochondrial Biogenesis Markers in Rat Skeletal Muscle. Int J Sports Med 37, 841–848.

Bishop P, Cureton K & Collins M (1987). Sex difference in muscular strength in equally-trained men and women. Ergonomics 30, 675–687.

Blencowe M, Chen X, Zhao Y, Itoh Y, McQuillen CN, Han Y, Shou BL, McClusky R, Reue K, Arnold AP & Yang X (2022). Relative contributions of sex hormones, sex chromosomes, and gonads to sex differences in tissue gene regulation. Genome Res 32, 807–824.

Blew RM, Lee VR, Farr JN, Schiferl DJ & Going SB (2014). Standardizing evaluation of pQCT image quality in the presence of subject movement: qualitative versus quantitative assessment. Calcif Tissue Int 94, 202–211.

Borg GA (1982). Psychophysical bases of perceived exertion. Med Sci Sports Exerc 14, 377–381.

Bowen TS, Schuler G & Adams V (2015). Skeletal muscle wasting in cachexia and sarcopenia: molecular pathophysiology and impact of exercise training. J Cachexia Sarcopenia Muscle 6, 197–207.

Callahan DM, Tourville TW, Miller MS, Hackett SB, Sharma H, Cruickshank NC, Slauterbeck JR, Savage PD, Ades PA, Maughan DW, Beynnon BD & Toth MJ (2015). Chronic disuse and skeletal muscle structure in older adults: sex-specific differences and relationships to contractile function. Am J Physiol-Cell Physiol 308, C932–C943.

Cervinka T, Giangregorio L, Sievanen H, Cheung AM & Craven BC (2018). Peripheral Quantitative Computed Tomography: Review of Evidence and Recommendations for Image Acquisition, Analysis, and Reporting, Among Individuals With Neurological Impairment. J Clin Densitom Off J Int Soc Clin Densitom 21, 563–582.

Chabi B, Ljubicic V, Menzies KJ, Huang JH, Saleem A & Hood DA (2008). Mitochondrial function and apoptotic susceptibility in aging skeletal muscle. Aging Cell 7, 2–12.

Chin ER, Olson EN, Richardson JA, Yang Q, Humphries C, Shelton JM, Wu H, Zhu W, Bassel-Duby R & Williams RS (1998). A calcineurin-dependent transcriptional pathway controls skeletal muscle fiber type. Genes Dev 12, 2499–2509.

Chinvattanachot G, Rivas D & Duque G (2024). Mechanisms of muscle cells alterations and regeneration decline during aging. Ageing Res Rev 102, 102589.

Cho E-J, Choi Y, Kim J, Bae JH, Cho J, Park D-H, Kang J-H, Yoon JH, Park E, Seo DY, Lee S & Kwak H-B (2021). Exercise Training Attenuates Ovariectomy-Induced Alterations in Skeletal Muscle Remodeling, Apoptotic Signaling, and Atrophy Signaling in Rat Skeletal Muscle. Int Neurourol J 25, S47–54.

Coggan AR, Spina RJ, King DS, Rogers MA, Brown M, Nemeth PM & Holloszy JO (1992). Histochemical and enzymatic comparison of the gastrocnemius muscle of young and elderly men and women. J Gerontol 47, B71–76.

Colley RC & Tremblay MS (2011). Moderate and vigorous physical activity intensity cut-points for the Actical accelerometer. J Sports Sci 29, 783–789.

Collins BC, Arpke RW, Larson AA, Baumann CW, Xie N, Cabelka CA, Nash NL, Juppi H-K, Laakkonen EK, Sipilä S, Kovanen V, Spangenburg EE, Kyba M & Lowe DA (2019). Estrogen Regulates the Satellite Cell Compartment in Females. Cell Rep 28, 368–381.e6.

Conesa A, Madrigal P, Tarazona S, Gomez-Cabrero D, Cervera A, McPherson A, Szcześniak MW, Gaffney DJ, Elo LL, Zhang X & Mortazavi A (2016). A survey of best practices for RNA-seq data analysis. Genome Biol 17, 13.

Critchlow AJ, Alexander SE, Hiam DS, Ferrucci L, Scott D & Lamon S (2025). Associations Between Female Sex Hormones and Skeletal Muscle Ageing: The Baltimore Longitudinal Study of Aging. J Cachexia Sarcopenia Muscle 16, e13786.

Critchlow AJ, Hiam D, Williams R, Scott D & Lamon S (2023). The role of estrogen in female skeletal muscle aging: A systematic review. Maturitas 178, 107844.

Cui J, Shen Y & Li R (2013). Estrogen synthesis and signaling pathways during aging: from periphery to brain. Trends Mol Med 19, 197–209.

Cuthbertson D, Smith K, Babraj J, Leese G, Waddell T, Atherton P, Wackerhage H, Taylor PM & Rennie MJ (2005). Anabolic signaling deficits underlie amino acid resistance of wasting, aging muscle. FASEB J Off Publ Fed Am Soc Exp Biol 19, 422–424.

D’Antona G (2003). The effect of ageing and immobilization on structure and function of human skeletal muscle fibres. J Physiol 552, 499–511.

De Jong JCBC, Attema BJ, Van Der Hoek MD, Verschuren L, Caspers MPM, Kleemann R, Van Der Leij FR, Van Den Hoek AM, Nieuwenhuizen AG & Keijer J (2023). Sex differences in skeletal muscle-aging trajectory: same processes, but with a different ranking. GeroScience 45, 2367–2386.

DeLuca DS, Levin JZ, Sivachenko A, Fennell T, Nazaire M-D, Williams C, Reich M, Winckler W & Getz G (2012). RNA-SeQC: RNA-seq metrics for quality control and process optimization. Bioinforma Oxf Engl 28, 1530–1532.

Ehmsen JT & Höke A (2020). Cellular and molecular features of neurogenic skeletal muscle atrophy. Exp Neurol 331, 113379.

Elliott-Sale KJ, Minahan CL, De Jonge XAKJ, Ackerman KE, Sipilä S, Constantini NW, Lebrun CM & Hackney AC (2021). Methodological Considerations for Studies in Sport and Exercise Science with Women as Participants: A Working Guide for Standards of Practice for Research on Women. Sports Med 51, 843–861.

Essén-Gustavsson B & Borges O (1986). Histochemical and metabolic characteristics of human skeletal muscle in relation to age. Acta Physiol Scand 126, 107–114.

Evans WJ, Phinney SD & Young VR (1982). Suction applied to a muscle biopsy maximizes sample size. Med Sci Sports Exerc 14, 101–102.

Fieldsend TW, O’Neill CR, Shrivastava A, Ogden HE, Dand N & Hughes SM (2025). Sexual dimorphism in human muscle ageing. 2025.01.06.25319958. Available at: https://www.medrxiv.org/content/10.1101/2025.01.06.25319958v1 [Accessed January 15, 2025].

Franceschi C & Campisi J (2014). Chronic Inflammation (Inflammaging) and Its Potential Contribution to Age-Associated Diseases. J Gerontol A Biol Sci Med Sci 69, S4–S9.

Frechette DM, Krishnamoorthy D, Adler BJ, Chan ME & Rubin CT (2015). Diminished satellite cells and elevated adipogenic gene expression in muscle as caused by ovariectomy are averted by low-magnitude mechanical signals. J Appl Physiol 119, 27–36.

Frederiksen H, Johannsen TH, Andersen SE, Albrethsen J, Landersoe SK, Petersen JH, Andersen AN, Vestergaard ET, Schorring ME, Linneberg A, Main KM, Andersson A-M & Juul A (2020). Sex-specific Estrogen Levels and Reference Intervals from Infancy to Late Adulthood Determined by LC-MS/MS. J Clin Endocrinol Metab 105, 754–768.

Gower BA & Nyman L (2000). Associations among Oral Estrogen Use, Free Testosterone Concentration, and Lean Body Mass among Postmenopausal Women.

Greising SM, Baltgalvis KA, Lowe DA & Warren GL (2009). Hormone Therapy and Skeletal Muscle Strength: A Meta-Analysis. J Gerontol A Biol Sci Med Sci 64A, 1071–1081.

Gumbiner BM (1996). Cell adhesion: the molecular basis of tissue architecture and morphogenesis. Cell 84, 345–357.

Haizlip KM, Harrison BC & Leinwand LA (2015). Sex-Based Differences in Skeletal Muscle Kinetics and Fiber-Type Composition. Physiology 30, 30–39.

Handelsman DJ, Sikaris K & Ly LP (2016). Estimating age-specific trends in circulating testosterone and sex hormone-binding globulin in males and females across the lifespan. Ann Clin Biochem Int J Lab Med 53, 377–384.

Hanks SC et al. (2025). Extensive differential gene expression and regulation by sex in human skeletal muscle. Cell Genomics100915.

Haynes EMK, Neubauer NA, Cornett KMD, O’Connor BP, Jones GR & Jakobi JM (2020). Age and sex-related decline of muscle strength across the adult lifespan: a scoping review of aggregated data. Appl Physiol Nutr Metab 45, 1185–1196.

He L, Khanal P, Morse CI, Williams A & Thomis M (2020). Associations of combined genetic and epigenetic scores with muscle size and muscle strength: a pilot study in older women. J Cachexia Sarcopenia Muscle 11, 1548–1561.

Heldring N, Pike A, Andersson S, Matthews J, Cheng G, Hartman J, Tujague M, Ström A, Treuter E, Warner M & Gustafsson J-Å (2007). Estrogen Receptors: How Do They Signal and What Are Their Targets. Physiol Rev 87, 905–931.

Hodgkinson K, Forrest LA, Vuong N, Garson K, Djordjevic B & Vanderhyden BC (2018). GREB1 is an estrogen receptor-regulated tumour promoter that is frequently expressed in ovarian cancer. Oncogene 37, 5873–5886.

Horwath O, Moberg M, Edman S, Philp A & Apró W (2025). Ageing leads to selective type II myofibre deterioration and denervation independent of reinnervative capacity in human skeletal muscle. Exp Physiol 110, 277–292.

Huang X, Chen M, Xiao Y, Zhu F, Chen L, Tian X & Hong L (2024). The influence of biological sex in human skeletal muscle transcriptome during ageing. Biogerontology 25, 461–478.

James JJ, Klevenow EA, Atkinson MA, Vosters EE, Bueckers EP, Quinn ME, Kindy SL, Mason AP, Nelson SK, Rainwater KAH, Taylor PV, Zippel EP & Hunter SK (2023). Underrepresentation of women in exercise science and physiology research is associated with authorship gender. J Appl Physiol 135, 932–942.

Jane FM & Davis SR (2014). A practitioner’s toolkit for managing the menopause. Climacteric J Int Menopause Soc 17, 564–579.

Janssen I, Heymsfield SB & Ross R (2002). Low Relative Skeletal Muscle Mass (Sarcopenia) in Older Persons Is Associated with Functional Impairment and Physical Disability. J Am Geriatr Soc 50, 889–896.

Javed AA, Mayhew AJ, Shea AK & Raina P (2019). Association Between Hormone Therapy and Muscle Mass in Postmenopausal Women: A Systematic Review and Meta-analysis. JAMA Netw Open 2, e1910154.

Juppi H-K, Sipilä S, Cronin NJ, Karvinen S, Karppinen JE, Tammelin TH, Aukee P, Kovanen V, Kujala UM & Laakkonen EK (2020). Role of Menopausal Transition and Physical Activity in Loss of Lean and Muscle Mass: A Follow-Up Study in Middle-Aged Finnish Women. J Clin Med 9, 1588.

Kadi F, Charifi N, Denis C & Lexell J (2004). Satellite cells and myonuclei in young and elderly women and men. Muscle Nerve 29, 120–127.

Kim TN & Choi KM (2013). Sarcopenia: Definition, Epidemiology, and Pathophysiology. J Bone Metab 20, 1.

Kitajima Y & Ono Y (2016). Estrogens maintain skeletal muscle and satellite cell functions. J Endocrinol 229, 267–275.

Klitgaard H, Zhou M, Schiaffino S, Betto R, Salviati G & Saltin B (1990). Ageing alters the myosin heavy chain composition of single fibres from human skeletal muscle. Acta Physiol Scand 140, 55–62.

Kong SH, Kim JH, Lee JH, Hong AR, Shin CS & Cho NH (2019). Dehydroepiandrosterone Sulfate and Free Testosterone but not Estradiol are Related to Muscle Strength and Bone Microarchitecture in Older Adults. Calcif Tissue Int 105, 285–293.

Kowarik A & Templ M (2016). Imputation with the R Package VIM. J Stat Softw 74, 1–16.

Lamon S, Soria M, Williams R, Critchlow A, Garnham A, Varshney A, Beillharz T, Hiam D & Ziemann M (2024). The transcriptomic signature of age and sex is not conserved in human primary myocytes.; DOI: 10.1101/2024.12.17.629041. Available at: http://biorxiv.org/lookup/doi/10.1101/2024.12.17.629041 [Accessed June 4, 2025].

Landen S, Hiam D, Voisin S, Jacques M, Lamon S & Eynon N (2023). Physiological and molecular sex differences in human skeletal muscle in response to exercise training. J Physiol 601, 419–434.

Larson AA, Baumann CW, Kyba M & Lowe DA (2020). Oestradiol affects skeletal muscle mass, strength and satellite cells following repeated injuries. Exp Physiol 105, 1700–1707.

Larsson L, Sjödin B & Karlsson J (1978). Histochemical and biochemical changes in human skeletal muscle with age in sedentary males, age 22–65 years. Acta Physiol Scand 103, 31– 39.

Lee C, Woods PC, Paluch AE & Miller MS (2024). Effects of age on human skeletal muscle: a systematic review and meta-analysis of myosin heavy chain isoform protein expression, fiber size, and distribution. Am J Physiol Cell Physiol 327, C1400–C1415.

Léger B, Derave W, De Bock K, Hespel P & Russell AP (2008). Human sarcopenia reveals an increase in SOCS-3 and myostatin and a reduced efficiency of Akt phosphorylation. Rejuvenation Res 11, 163–175B.

Lim J-Y, Choi SJ, Widrick JJ, Phillips EM & Frontera WR (2019). Passive force and viscoelastic properties of single fibers in human aging muscles. Eur J Appl Physiol 119, 2339–2348.

Mansouri A, Muller FL, Liu Y, Ng R, Faulkner J, Hamilton M, Richardson A, Huang T-T, Epstein CJ & Van Remmen H (2006). Alterations in mitochondrial function, hydrogen peroxide release and oxidative damage in mouse hind-limb skeletal muscle during aging. Mech Ageing Dev 127, 298–306.

McClung JM, Davis JM, Wilson MA, Goldsmith EC & Carson JA (2006). Estrogen status and skeletal muscle recovery from disuse atrophy. J Appl Physiol 100, 2012–2023.

McGlory C, van Vliet S, Stokes T, Mittendorfer B & Phillips SM (2019). The impact of exercise and nutrition on the regulation of skeletal muscle mass. J Physiol 597, 1251–1258.

Miller MS, Bedrin NG, Callahan DM, Previs MJ, Jennings ME, Ades PA, Maughan DW, Palmer BM & Toth MJ (2013). Age-related slowing of myosin actin cross-bridge kinetics is sex specific and predicts decrements in whole skeletal muscle performance in humans. J Appl Physiol 115, 1004–1014.

Noehren B, Hardy PA, Andersen A, Brightwell CR, Fry JL, Vandsburger MH, Thompson KL & Fry CS (2021). T1ρ imaging as a non-invasive assessment of collagen remodelling and organization in human skeletal muscle after ligamentous injury. J Physiol 599, 5229–5242.

Nyberg M, Egelund J, Mandrup CM, Andersen CB, Hansen KMBE, Hergel IF, Valbak-Andersen N, Frikke-Schmidt R, Stallknecht B, Bangsbo J & Hellsten Y (2017). Leg vascular and skeletal muscle mitochondrial adaptations to aerobic high-intensity exercise training are enhanced in the early postmenopausal phase. J Physiol 595, 2969–2983.

O’Bryan SJ, Critchlow A, Fuchs CJ, Hiam D & Lamon S (2025). The contribution of age and sex hormones to female neuromuscular function across the adult lifespan. J Physiol; DOI: 10.1113/JP287496.

Oliva M et al. (2020). The impact of sex on gene expression across human tissues. Science 369, eaba3066.

Park Y-M, Jankowski CM, Ozemek C, Hildreth KL, Kohrt WM & Moreau KL (2020). Appendicular lean mass is lower in late compared with early perimenopausal women: potential role of FSH. J Appl Physiol 128, 1373–1380.

Pataky MW, Dasari S, Michie KL, Sevits KJ, Kumar AA, Klaus KA, Heppelmann CJ, Robinson MM, Carter RE, Lanza IR & Nair KS (2023). Impact of biological sex and sex hormones on molecular signatures of skeletal muscle at rest and in response to distinct exercise training modes. Cell Metab 35, 1996–2010.e6.

Pellegrino A, Tiidus PM & Vandenboom R (2022). Mechanisms of Estrogen Influence on Skeletal Muscle: Mass, Regeneration, and Mitochondrial Function. Sports Med 52, 2853–2869.

Pöllänen E, Kangas R, Horttanainen M, Niskala P, Kaprio J, Butler-Browne G, Mouly V, Sipilä S & Kovanen V (2015). Intramuscular sex steroid hormones are associated with skeletal muscle strength and power in women with different hormonal status. Aging Cell 14, 236–248.

Pöllänen E, Sipilä S, Alen M, Ronkainen PHA, Ankarberg-Lindgren C, Puolakka J, Suominen H, Hämäläinen E, Turpeinen U, Konttinen YT & Kovanen V (2011). Differential influence of peripheral and systemic sex steroids on skeletal muscle quality in pre- and postmenopausal women. Aging Cell 10, 650–660.

Puig RR, Boddie P, Khan A, Castro-Mondragon JA & Mathelier A (2021). UniBind: maps of high-confidence direct TF-DNA interactions across nine species. BMC Genomics 22, 482.

Rariy CM, Ratcliffe SJ, Weinstein R, Bhasin S, Blackman MR, Cauley JA, Robbins J, Zmuda JM, Harris TB & Cappola AR (2011). Higher serum free testosterone concentration in older women is associated with greater bone mineral density, lean body mass, and total fat mass: the cardiovascular health study. J Clin Endocrinol Metab 96, 989–996.

Sala D & Sacco A (2016). Signal transducer and activator of transcription 3 signaling as a potential target to treat muscle wasting diseases: Curr Opin Clin Nutr Metab Care1.

Sato T, Akatsuka H, Kito K, Tokoro Y, Tauchi H & Kato K (1984). Age changes in size and number of muscle fibers in human minor pectoral muscle. Mech Ageing Dev 28, 99–109.

Schindelin J, Arganda-Carreras I, Frise E, Kaynig V, Longair M, Pietzsch T, Preibisch S, Rueden C, Saalfeld S, Schmid B, Tinevez J-Y, White DJ, Hartenstein V, Eliceiri K, Tomancak P & Cardona A (2012). Fiji: an open-source platform for biological-image analysis. Nat Methods 9, 676–682.

Shock NW (1984). Normal Human Aging: The Baltimore Longitudinal Study of Aging. Superintendent of Documents, U. Available at: https://eric.ed.gov/?id=ED292030 [Accessed June 4, 2025].

Sipilä S, Heikkinen E, Cheng S, Suominen H, Saari P, Kovanen V, Alén M & Rantanen T (2006). Endogenous Hormones, Muscle Strength, and Risk of Fall-Related Fractures in Older Women. J Gerontol A Biol Sci Med Sci 61, 92–96.

Sishi BJN & Engelbrecht A-M (2011). Tumor necrosis factor alpha (TNF-α) inactivates the PI3-kinase/PKB pathway and induces atrophy and apoptosis in L6 myotubes. Cytokine 54, 173– 184.

St-Jean-Pelletier F, Pion CH, Leduc-Gaudet J, Sgarioto N, Zovilé I, Barbat-Artigas S, Reynaud O, Alkaterji F, Lemieux FC, Grenon A, Gaudreau P, Hepple RT, Chevalier S, Belanger M, Morais JA, Aubertin-Leheudre M & Gouspillou G (2017). The impact of ageing, physical activity, and pre-frailty on skeletal muscle phenotype, mitochondrial content, and intramyocellular lipids in men. J Cachexia Sarcopenia Muscle 8, 213–228.

Swislocki ALM & Eisenberg ML (2024). A Review on Testosterone: Estradiol Ratio—Does It Matter, How Do You Measure It, and Can You Optimize It? World J Mens Health; DOI: 10.5534/wjmh.240029.

Tomazin K, Verges S, Decorte N, Oulerich A, Maffiuletti NA & Millet GY (2011). Fat tissue alters quadriceps response to femoral nerve magnetic stimulation. Clin Neurophysiol 122, 842– 847.

Van De Casteele F, Van Thienen R, Horwath O, Apró W, Van Der Stede T, Moberg M, Lievens E & Derave W (2024). Does one biopsy cut it? Revisiting human muscle fiber type composition variability using repeated biopsies in the vastus lateralis and gastrocnemius medialis. J Appl Physiol 137, 1341–1353.

Verdijk LB, Koopman R, Schaart G & Meijer K (2007). Satellite cell content is specifically reduced in type II skeletal muscle fibers in the elderly. Endocrinol Metab.

Wang X, Park J, Susztak K, Zhang NR & Li M (2019). Bulk tissue cell type deconvolution with multi-subject single-cell expression reference. Nat Commun 10, 380.

Wiik A, Ekman M, Johansson O, Jansson E & Esbjörnsson M (2009). Expression of both oestrogen receptor alpha and beta in human skeletal muscle tissue. Histochem Cell Biol 131, 181–189.

Wilson JM, Loenneke JP, Jo E, Wilson GJ, Zourdos MC & Kim J-S (2012). The Effects of Endurance, Strength, and Power Training on Muscle Fiber Type Shifting. J Strength Cond Res 26, 1724–1729.

Wohlgemuth SE, Seo AY, Marzetti E, Lees HA & Leeuwenburgh C (2010). Skeletal muscle autophagy and apoptosis during aging: Effects of calorie restriction and life-long exercise. Exp Gerontol 45, 138–148.

Wood N, Critchlow A, Cheng CW, Straw S, Hendrickse PW, Pereira MG, Wheatcroft SB, Egginton S, Witte KK, Roberts LD & Bowen TS (n.d.). Sex Differences in Skeletal Muscle Pathology in Patients With Heart Failure and Reduced Ejection Fraction. Circ Heart Fail 0, e011471.

Wood TM, Maddalozzo, Gianni F. & Harter RA (2002). Accuracy of Seven Equations for Predicting 1-RM Performance of Apparently Healthy, Sedentary Older Adults. Meas Phys Educ Exerc Sci 6, 67–94.

Xu H & Van Remmen H (2021). The SarcoEndoplasmic Reticulum Calcium ATPase (SERCA) pump: a potential target for intervention in aging and skeletal muscle pathologies. Skelet Muscle 11, 25.

